# The Effects of Extreme Heat on Mental Disorder Admissions in Hlabisa, South Africa: A 13-Year Time-Series Analysis

**DOI:** 10.64898/2026.06.18.26355969

**Authors:** Elisabeth Lindner, Frank Tanser, Kobus Herbst, Thobeka Mngomezulu, Dickman Gareta, Till Bärnighausen, Gillian F. Black, Marlies H Craig, Vasco Cumbe, Sammy Khagayi, Frederick Murunga Wekesah, Alberto Gabriel Muanido, Chanelle Mulopo, Sithembiso Ndlovu, Evans Omondi, Flavian Otieno, Henry Owoko, Ali Sie, Mareca Sithole, Moustapha Tall, Nafissatou Traoré, Astrid Treffry-Goatley, G. Nduku Wambua, Prasad Liyanage, Collins Iwuji, Aditi Bunker

## Abstract

**Background:** Extreme heat is increasingly linked to adverse mental health outcomes, yet empirical evidence from sub-Saharan Africa is absent. Such data are critical for health system planning and climate adaptation in one of the world’s most heat-vulnerable regions.

**Objectives:** We aimed to estimate the immediate and delayed effects of daily maximum temperature on mental disorder-related hospital admissions in a rural South African district over 13 years.

**Methods:** We conducted a time-series analysis of daily mental disorder admissions at Hlabisa Hospital, South Africa, from 2011 to 2023. Daily maximum temperatures were obtained from ECMWF ERA5 reanalysis and validated with local weather station data. Using distributed lag non-linear quasi-Poisson models, we estimated the association between extreme heat (95th, 97.5th, 99th percentiles of the daily maximum temperature distribution) and relative risk of mental disorder admissions over a 0-21-day lag. Analyses were stratified by sex, age (<60 vs. ≥60 years), and diagnosis (schizophrenia vs. other mental disorders).

**Results:** Between 2011 and 2023, 1760 mental disorder admissions were recorded. Extreme heat at the 97.5th percentile (35.74°C) was associated with an immediate increase in admissions (lag 0: RR=1.10, 95 % CI: 1.00–1.20; lag 1: RR=1.08, 95% CI: 1.00-1.17). Subgroup analyses revealed heterogeneous patterns, with early increases among females and individuals with diagnoses other than schizophrenia, delayed increases among males and those aged <60 years, and delayed decreases among females and older adults. No significant association was observed for schizophrenia.

**Discussion:** To our knowledge, this study provides the first empirical evidence from South Africa that extreme heat is associated with increased hospital admissions for mental disorders, with immediate and delayed effects across population subgroups. These findings highlight the need to integrate mental health into climate adaptation and public health strategies through strengthened surveillance, targeted protection of vulnerable groups, and enhanced health system preparedness for extreme heat.

## Introduction

Mental disorders represent a significant and escalating challenge to global public health, ranking seventh among the leading causes of disease burden worldwide.^1^ In 2019, an estimated 970 million people were living with a mental disorder globally.^1^ Concurrently, climate change—particularly global warming and the increasing frequency of extreme weather events—is posing an escalating threat to public health, with Sub-Saharan Africa projected to warm at a higher rate than the global average.^2^ A growing body of evidence links extreme temperature to adverse mental health outcomes.^2^ Evidence from sub-Saharan Africa on the association between extreme heat and mental disorders, however, remains limited. This gap is particularly important in rural settings, where social, economic, and health system vulnerabilities directly shape mental health outcomes and may further modulate both exposure to extreme heat and access to care.^3,4^

Amid high levels of poverty, income inequality, and a substantial burden of HIV, the South African population is particularly vulnerable to mental illness.^4–7^ While recent data on the prevalence of mental disorders in South Africa are limited, a 2009 study reported a lifetime prevalence of 30.3% for any mental disorder.^8^ More recent estimates suggest that over a quarter of South Africans experience moderate to severe symptoms of probable depression, with prevalence ranging from 14.7 to 38.8%. ^9^

In South Africa, each 1°C increase in daily maximum temperature has been associated with a 1.5% increase in homicide risk, with odds approximately 18% higher at temperatures between 30 and 35°C compared with those below 20°C.^10^ Violent crime more broadly has been reported to be around 50% more frequent on the hottest days of the year than on the coldest days.^11^ High ambient temperatures have also been linked to increased suicide risk; one multi-country study reported an approximately 79% higher risk at the maximum suicide temperature (26.4°C) compared with the minimum (7.6°C) in South Africa, although this estimate should be interpreted cautiously given limitations in suicide mortality data, including low statistical power due to small sample sizes and the frequent underreporting or misclassification of violent deaths in national vital statistics.^12^

Projections indicate that South Africa will experience substantial increases in mean annual temperature alongside more frequent and prolonged periods of extreme heat, underscoring the urgency of examining their potential mental health impacts.^13^ Despite these risks, little is known about the association between heat and mental health outcomes, particularly in rural South Africa. To address this gap, our current study provides, to our knowledge, the first investigation of the association between extreme temperatures and hospital admissions related to mental disorder in South Africa. Using a time series design, we examine the lag-exposure-response relationship between daily maximum temperature and hospital admissions in Hlabisa, KwaZulu-Natal, South Africa.

## Methods

### Study area and data collection

We utilized data from the Africa Health Research Institute (AHRI) Hospital Information System (HIS), comprising records of individuals admitted to and discharged from Hlabisa Hospital between 1 January 2011 and 30 December 2023.^14^ The hospital is located in the uMkhanyakude district, northwest of the *Hluhluwe-Umfolozi* Game Reserve in KwaZulu-Natal Province, South Africa (Figure 1). It serves communities in Hlabisa, Mtubatuba, and parts of the Big Five Hlabisa municipality and is supported by 17 fixed clinics. The HIS dataset includes unique patient identifiers, date of birth, sex, and admission and discharge diagnoses. We selected records where the primary discharge diagnosis was classified as a mental disorder, defined using the International Classification of Disease Tenth Revision (ICD-10) codes F00-F99 (mental and behavioral disorders) and X60-X84 (intentional self-harm). This study received ethics approval from the University of KwaZulu-Natal Biomedical Research Ethics Committee (BREC) (reference number: BREC/00007752/2024), Comite d Ethique pour la Recherche en Sante (CERS) for Burkina Faso (reference number: 2024-12-376), Amref Ethics and Scientific Review Committee (ESRC) for Kenya (reference number: ESRC P1792/2024), and Comite Nacional de Bioetica para Saudes (CNBS) for Mozambique (reference number: 309/CNBS/25).

**Figure 1.**
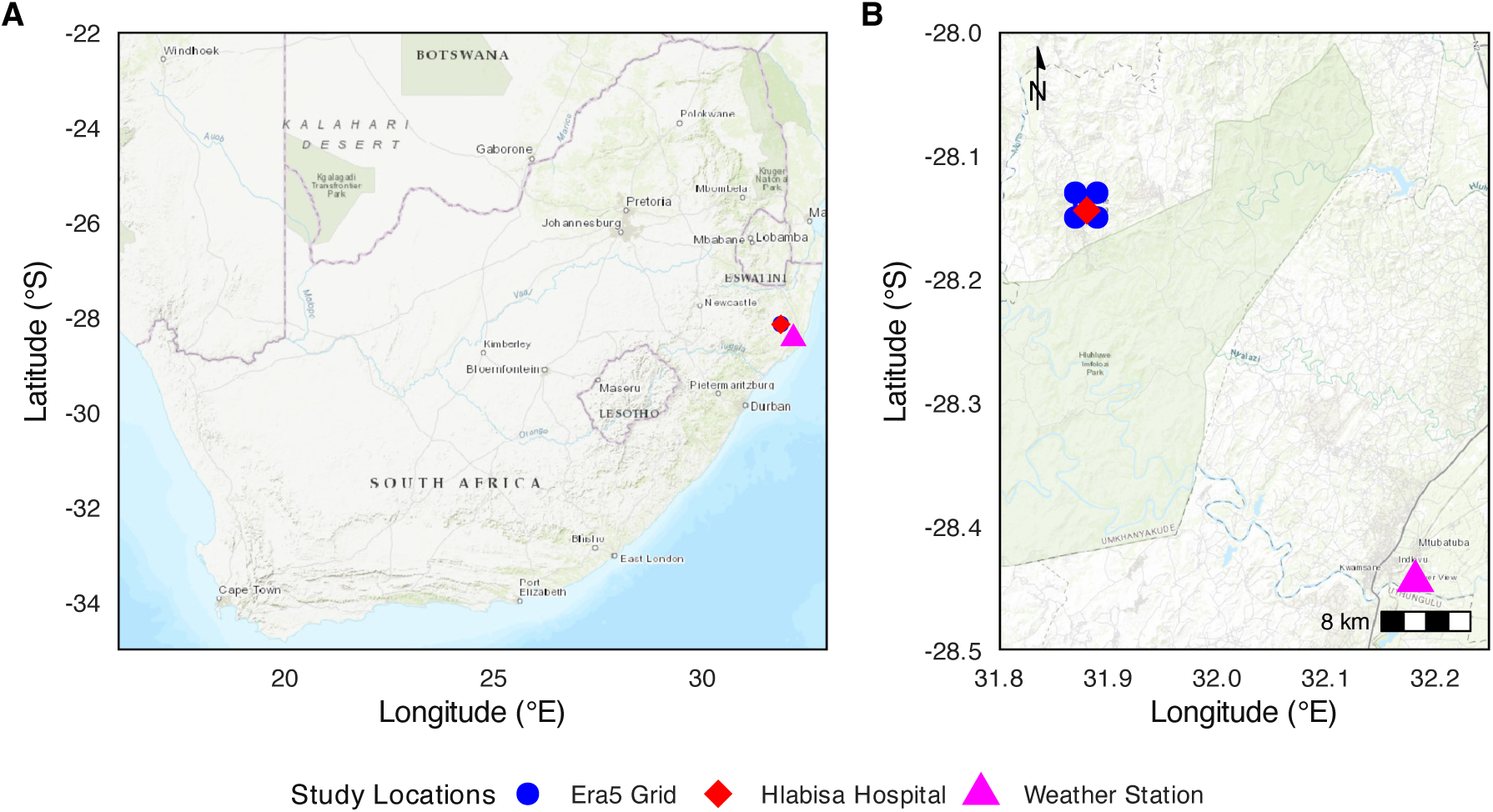
Study locations in South Africa and zoomed-in grid around Hlabisa. (A) Map of South Africa showing the study locations: ERA5 grid points (blue circles), Hlabisa Hospital (red diamond), and the Riverview weather station (magenta triangle). (B) Zoomed-in view of the study area around Hlabisa with ERA5 grid points (blue circles), Hlabisa Hospital (red diamond), and the Riverview weather station (magenta triangle).

### Exposure variable data

We obtained hourly meteorological data from the fifth-generation European Reanalysis (ERA5), produced by the European Centre for Medium Range Weather Forecasts (ECMWF) under the Copernicus program.^15^ Temperature at 2 meters above ground level was extracted at a spatial resolution of 0.25° x 0.25°, and daily maximum temperature was derived from hourly values. Meteorological conditions were represented using data from the four grid points closest to Hlabisa Hospital (16.60 – 20.57 km from the site). To ensure temporal consistency and avoid irregularities in the daily time series, February 29^th^ was excluded from all leap years.

## Statistical analysis

We applied distributed lag non-linear models (DLNM) to capture the delayed and non-linear effects of temperature on mental disorder hospitalization.^16,17^ These models were used to estimate the association between daily maximum temperature and the relative risk (RR) of hospital admissions for mental disorder, considering both immediate (lag 0) and delayed effects (lag *l*) up to 21 days after daily maximum temperature exposure.^16^ This 21-day lag window was selected based on previous literature identifying that heat can have delayed effects on mental health, extending beyond the immediate exposure period.^16^

We modeled daily counts of hospital admissions for mental disorders *Y_t_* using a quasi-Poisson generalized linear model to account for overdispersion combined with DLNM:

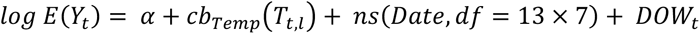

where *E*(*Y_t_*) is the estimated daily number of admissions on day *t* and *cb_Temp_* is the cross-basis function for maximum daily temperature *T* over lags *l* = 0 to 21 days. The term *ns*(*Date*, *df* = 13 × 7) represents a natural spline for long-term trend and seasonality with 7 df per year over 13 years, as suggested in the literature.^16^ *DOW_t_* is a categorical day-of-week term.^16^ Each cross-basis function consists of a natural cubic exposure-response spline and a natural cubic lag-response spline over the fixed 21-day lag window.

To identify optimal knot placements for cross-basis splines, we conducted a parallelized grid search over both the exposure-response and lag-response dimensions of the cross-basis: Temperature exposure knots: all ordered pairs (*k*_1_, *k*_2_) with *k*_1_ ∈ {26, 27 28, 29, 30} °C and *k*_2_ ∈ {27 28, 29, 30,31,32,33} °C, subject to *k*_1_ > *k*_2_. Temperature lag knots: tested on day 6, day 14, or both days 7 and 14. All combinations of exposure- and lag-knots were fitted, and models were ranked using the quasi-Akaike Information Criterion (qAIC), separately for ERA5 and local weather station data (WS).^16,17^ The specification with the lowest qAIC was selected, with temperature knots at 30°C (ERA5: 73.27^th^ percentile, WS: 75.32nd percentile) and 33°C (ERA5: 90.49^th^ percentile, WS: 92.01st percentile) and a lag knot at 14 days (see Supplementary Information Table S1-S2 for the ten highest qAIC configurations). Using the final model, we estimated lag-specific and cumulative relative risks (RR) of hospital admission for mental disorders relative to the median daily maximum temperature. We report RRs at extreme heat, defined as the 95^th^ (ERA5: 34.22°C, WS: 33.90°C), 97·5^th^ (ERA5: 35.74°C, WS: 35.30°C) and 99^th^ (ERA5: 37.16°C, WS: 37.26°C) percentiles of daily maximum temperature. Lag-specific percentiles were rounded to the nearest 0.5°C. Estimates are presented with 95% confidence intervals (CI) and calculated for both individual and cumulative lagged days of exposure.^16^ We further conducted stratified analyses by sex, age (<60 and ≥60), and mental health diagnoses (schizophrenia vs other disorders) using ERA5 data. All statistical analyses were performed in R (version 4.4.2) using the DLNM and splines packages.^18^

### Sensitivity analysis

We repeated the main analysis using data from the local Riverview weather station (44.55 km from Hlabisa hospital; r=0.94 with ERA5; 0.49% of missing values imputed using ERA5) to assess robustness to the ERA5 data. To evaluate potential over-adjustment for seasonality, we additionally extended the grid search to identify optimal knot placement, as described in the statistical analysis section, across a range of seasonal degrees of freedom (35 to 105 in steps of 7) and reran the main model using the optimal specification.

## Results

### Descriptive results

Between 1 January 2011 and 30 December 2023, (excluding February 29 in leap years) 90748 admissions were recorded at Hlabisa Hospital. Of these, 1760 (1.94%) had a primary discharge diagnosis related to mental disorders (Figures 2-3). Among these admissions, 836 (47.50 %) were female and 924 (52.50 %) were male. Admissions varied across age groups, with 97 patients (5.51%) aged 0-17 years, 1086 (61.70%) aged 18-39, 421 (23.92%) aged 40-59 years, and 156 (8.86%) aged ≥ 60 years (Figure 3, Table 1).

**Figure 2.**
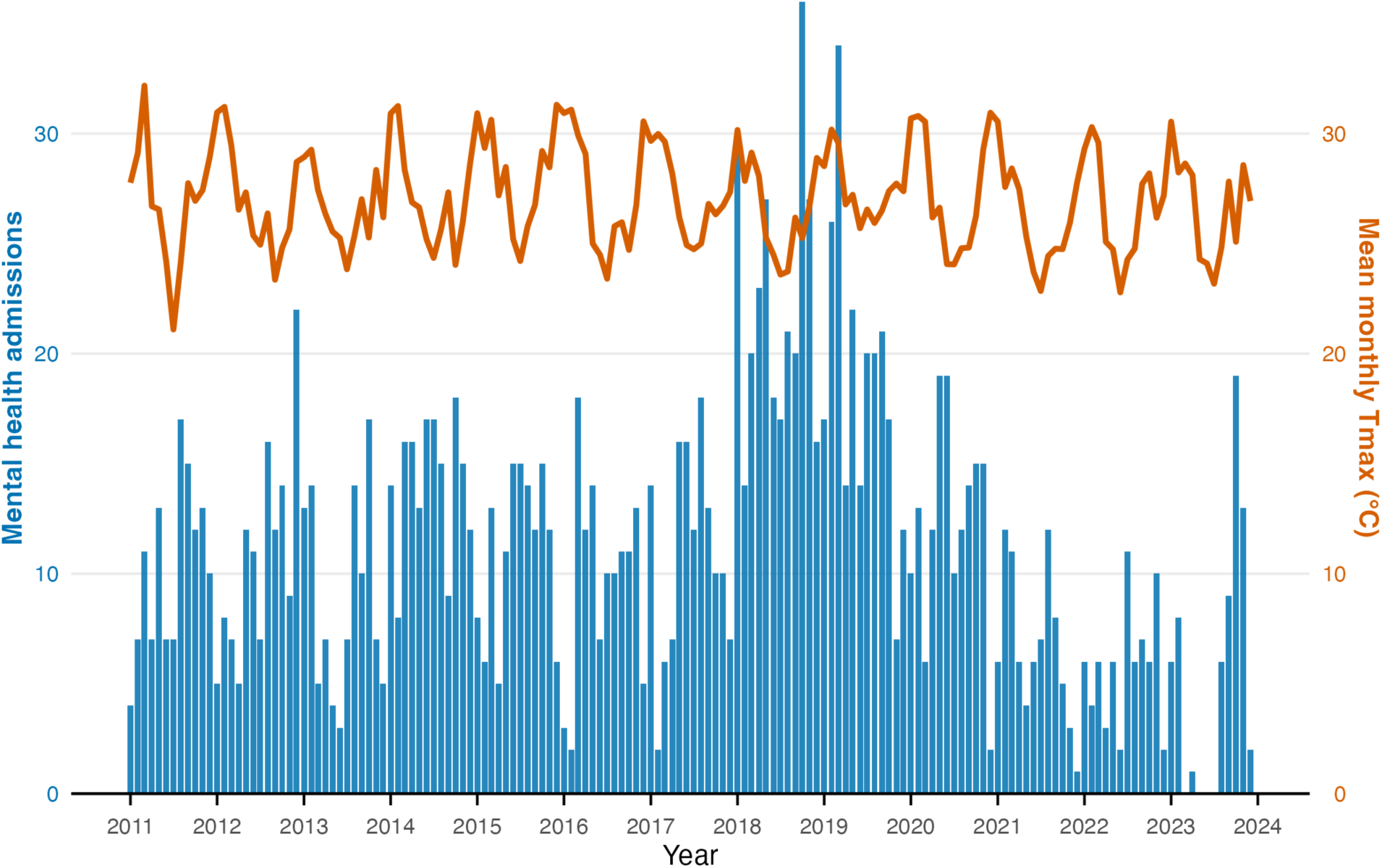
Monthly mental disorder admissions and maximum temperature, 2011-2023. Blue bars represent total monthly mental disorder admissions. The orange line indicates the mean monthly daily maximum temperature in °C.

**Figure 3.**
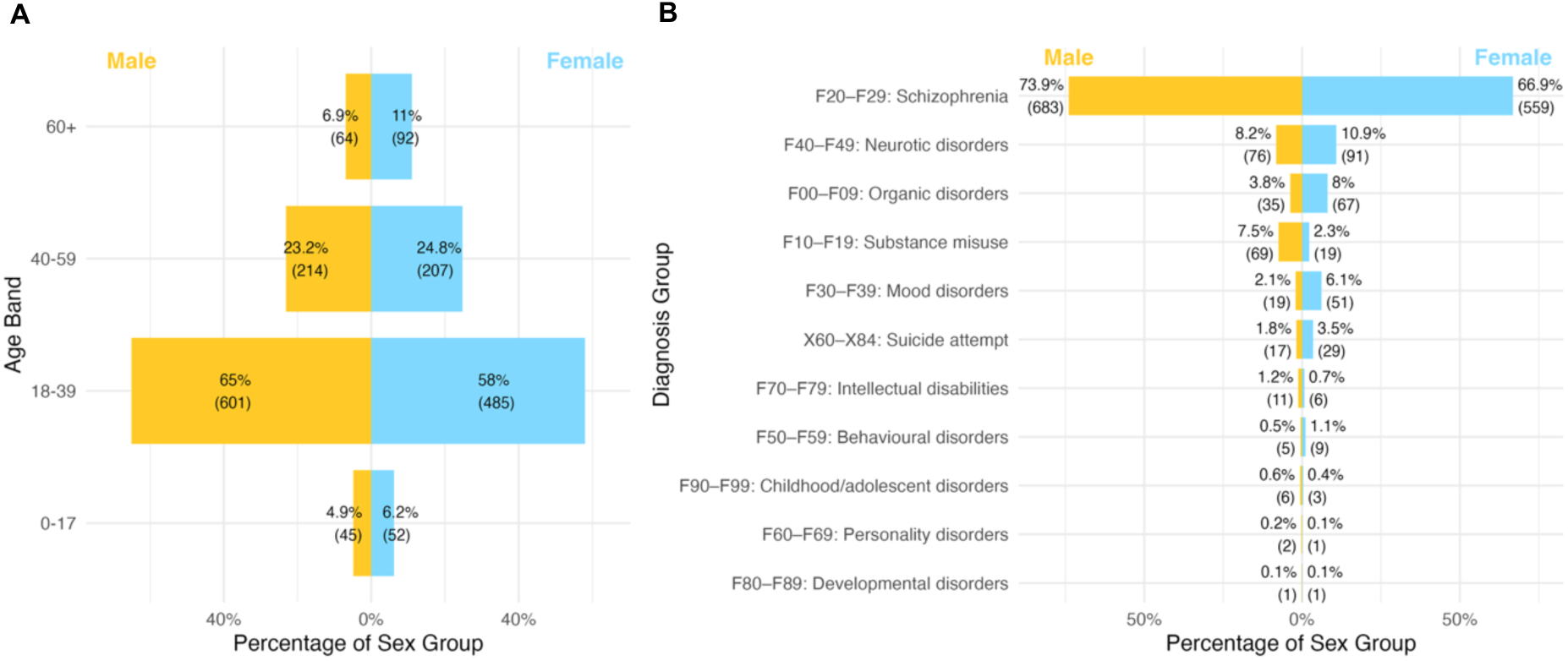
Mental disorder admissions by age band and sex (A) and by diagnosis group and sex (B). Data are shown as percentages with absolute numbers in brackets. Males are shown in yellow and females in blue.

**Table 1.** Descriptive statistics for mental disorder admissions to Hlabisa Hospital (South Africa) according to primary discharge diagnosis, 2011-2023.

| Characteristic | N | % |
| --- | --- | --- |
| **Total mental health admissions** | 1760 |  |
| **Proportion of all hospital admissions** |  | 1.94% |
| **Age distribution** |  |  |
| 0-17 years | 97 | 5.51 |
| 18-39 years | 1086 | 61.70 |
| 40-59 years | 421 | 23.92 |
| 60+ years | 156 | 8.86 |
| **Sex** |  |  |
| Male | 924 | 52.50 |
| Female | 836 | 47.50 |
| **Primary discharge diagnosis (ICD-10)** |  |  |
| F20–F29: Schizophrenia | 1242 | 70.57 |
| F40–F49: Neurotic disorders | 167 | 9.49 |
| F00–F09: Organic disorders | 102 | 5.80 |
| F10–F19: Substance misuse | 88 | 5.00 |
| F30–F39: Mood disorders | 70 | 3.98 |
| X60–X84: Suicide attempt | 46 | 2.61 |
| F70–F79: Intellectual disabilities | 17 | 0.97 |
| F50–F59: Behavioral disorders | 14 | 0.80 |
| F90–F99: Childhood/adolescent disorders | 9 | 0.51 |
| F60–F69: Personality disorders | 3 | 0.17 |
| F80–F89: Developmental disorders | 2 | 0.11 |

\* Exclusion: February 29 leap day admissions removed.

The mean daily temperature over the study period was 21.25°C (standard deviation (SD) 3.62°C). The mean daily maximum temperature was 27.00°C (SD 4.54°C) ranging from 12.37°C to 42.37°C, with a median of 27.03°C.

### Association between daily maximum temperature and mental disorder-related admissions

We found lag-dependent associations between daily maximum temperature and the RR of mental disorder admissions, using the median daily maximum temperature of 27.03°C as the reference (Figures 4-5). Temperatures at the 97.5^th^ percentile (35.74°C), were associated with an immediate increase in risk at lag 0 (RR = 1.10, 95% CI: 1.00 – 1.20) and lag 1 (RR = 1.08, 95% CI: 1.00-1.17) following the exposure.

**Figure 4.**
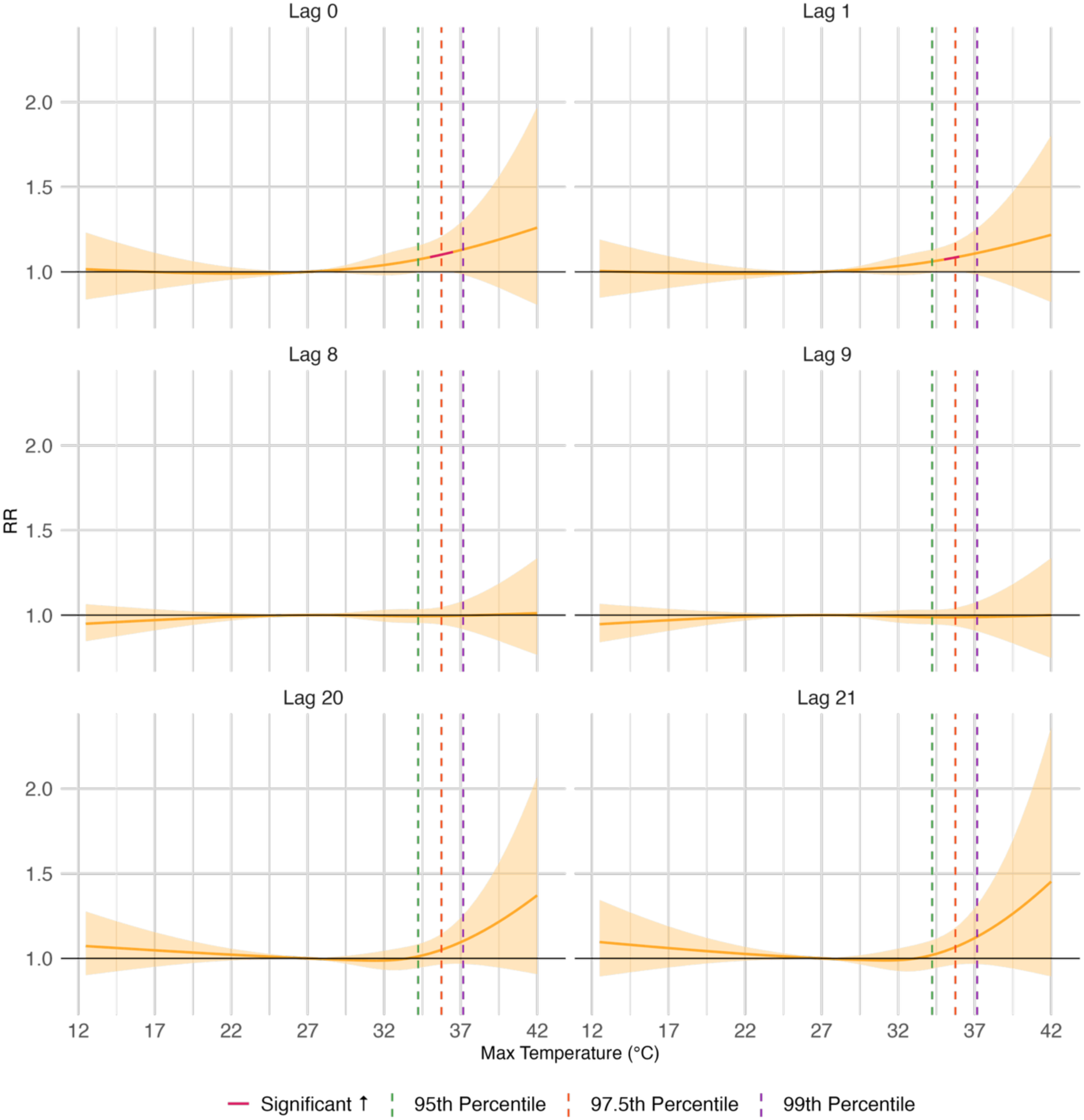
Lag-specific exposure-response curves of maximum daily temperature (°C) and relative risk of admission for mental disorders (relative to the median temperature), based on ERA5 temperature data. Orange and red response curve lines correspond to non-significant and significant increased relative risk (RR), respectively. Green, dark orange, and purple dotted vertical lines mark the 95th, 97·5th and 99th temperature percentile, respectively. Shaded areas represent 95% confidence intervals.

**Figure 5.**
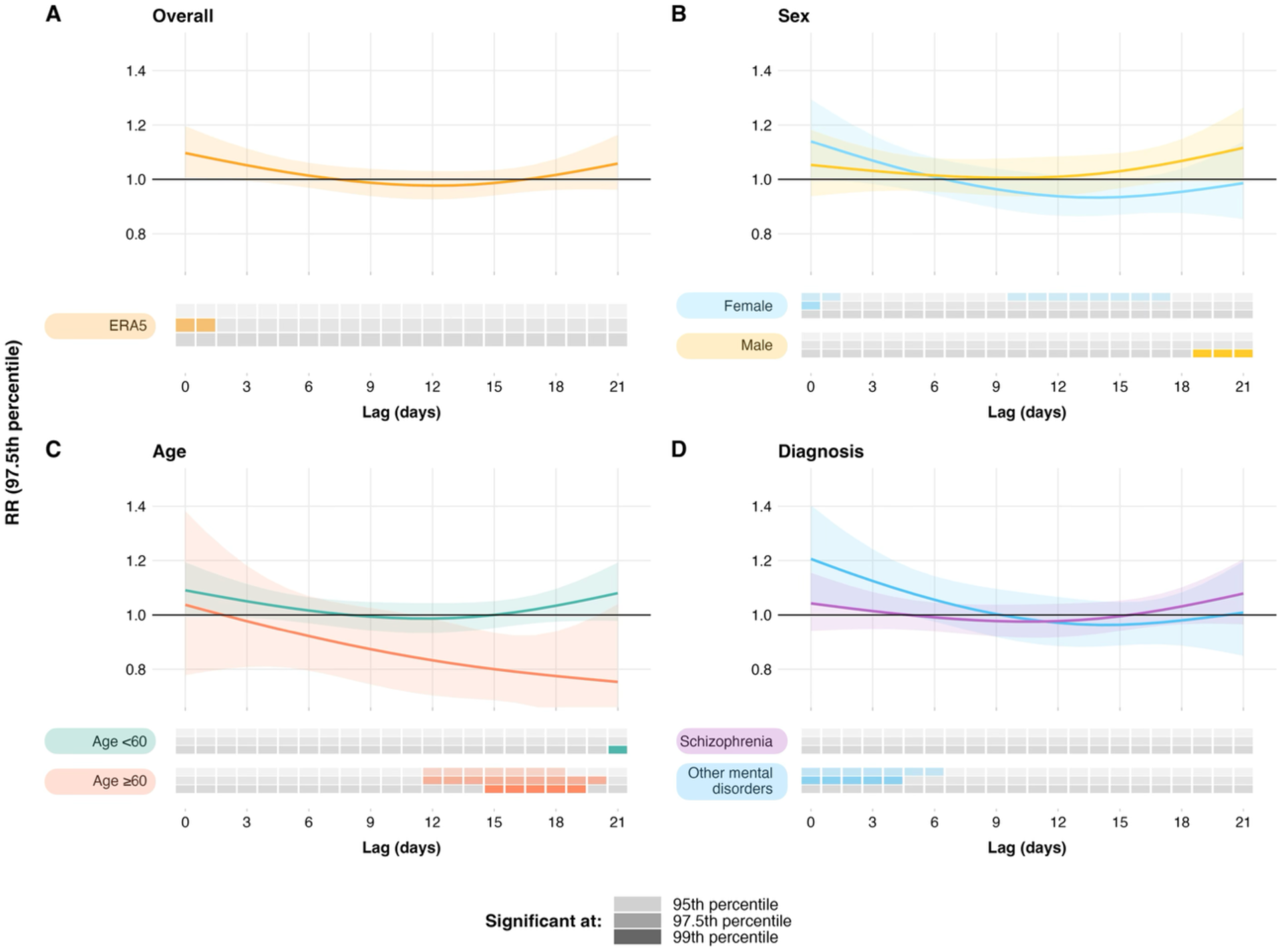
Lag-specific associations between extreme heat and mental disorder related hospital admissions by subgroup. (A) Overall population; (B) Sex; (C) Age; and (D) Diagnosis. Data are presented as relative risks (RR) at the 97·5th percentile of daily maximum temperature distribution compared with the median. Upper panels show fitted RRs (solid lines) with 95% CIs (shaded areas). Lower panels indicate lag-specific statistical significance across three heat intensity thresholds, represented as stacked vertical positions within each group: 95th percentile (top slot, light shading), 97.5th percentile (middle slot, medium shading), and 99th percentile (bottom slot, solid shading).

### Subgroup Analyses

Stratified analyses revealed subgroup-specific immediate and delayed effects (Figure 5; also see Supplementary Information). Among females, we observed a significant increase in the RR of mental disorder admissions at immediate lags (0-1) peaking at lag 0 at the 97.5^th^ percentile (35.74°C; RR = 1.14, 95% CI: 1.00-1.30). Across delayed lags (10-17), the RR decreased at the 95^th^ percentile, reaching a minimum at lag 14 (RR = 0.92, 95% CI: 0.86-0.98). The cumulative RR was significantly elevated over lag 0-3 at the 95^th^ percentile (34.22°C; RR = 1.44, 95% CI: 1.02-2.04).

In contrast, males showed no significant immediate-lag association. Instead, RR increased at delayed lags (19-21) at the 99^th^ percentile, peaking at lag 21 (37.16°C; RR = 1.23, 95% CI: 1.02-1.47).

In individuals aged <60 years, we observed an increase in RR of all mental disorder admissions at lag 21 (99^th^ percentile, RR = 1.16, 95% CI: 1.00-1.35) whereas those aged ≥60 years showed decreased RR at lags 12-20. We observed the lowest RR in mental disorder admissions in people aged ≥60 at lag 19 at the 99^th^ percentile (37.16°C; RR = 0.67, 95% CI: 0.45-0.98), and a reduced cumulative RR for lag 0-21 (99^th^ percentile, RR = 0.00, 95% CI: 0.00-0.56; 97.5^th^ percentile, RR = 0.03, 95% CI: 0.00-0.58).

We found no statistically significant associations between heat exposure and schizophrenia admissions, which we assessed due to 70.57% of all hospital admissions being attributable to schizophrenia diagnoses. In contrast, all other mental health diagnoses, excluding schizophrenia, showed a significant increase in RR at early lags (0-6) peaking at lag 0 at the 95^th^ percentile (34.22°C; RR = 1.23, 95% CI: 1.09-1.40). Significant increases were also observed for cumulative lag windows 0-3 (maximum RR = 1.99, 95% CI: 1.32-2.99, 95^th^ percentile), 0-7 (maximum RR = 2.77, 95% CI: 1.48-5.17, 95^th^ percentile) and 0-14 (RR = 2.76, 95% CI: 1.11-6.85, 95^th^ percentile).

### Sensitivity Analyses

In our sensitivity analysis, weather station data showed comparable temperature distribution with the ERA5 dataset (r = 0.94, mean daily maximum temperature at 27.17°C, SD 4.15°C, ranging 12.8°C to 43.3°C, median 27.1°C). Results on the effect of temperature on overall mental disorder admissions did not vary greatly with the use of weather station data replacing ERA5 data. We observed a significant increase in the immediate RR of all mental disorder admissions at lag 0 for the 97·5^th^ percentile (35.30°C; RR = 1.11, 95% CI: 1.01-1.22) and 99^th^ percentile (37.26°C; RR = 1.18, 95% CI: 1.00-1.39), and at lag 1 for the 97·5^th^ percentile (35.30°C; RR = 1.09, 95% CI: 1.00-1.19).

When the seasonality term was jointly optimized alongside exposure and lag knots, the extended grid search identified 49 df (3.77 df per year, Supplementary Information Table S3). Under this specification, a significant immediate increase in the RR was observed at the 95th percentile at lag 0 (34.22°C; RR = 1.07, 95% CI: 1.01 – 1.14) and lag 1 (RR = 1.06, 95% CI: 1.00-1.11) with a significant cumulative increased RR over lags 0-3 at the 95th percentile (34.22°C; RR = 1.23, 95% CI: 1.00 – 1.51), confirming that the primary findings are not attributable to over-adjustments of the seasonality term.

## Discussion

Our current study is the first, to our knowledge, that has shown an association between extreme heat exposure and increased risk of immediate hospital admissions from mental disorders in Hlabisa, South Africa. The immediate risk was greatest among females and individuals with non-schizophrenia diagnoses, whereas males and those aged <60 years exhibited delayed increases in risk at longer lags. Conversely, females and older adults (≥60 years) showed decreased risk at delayed lags. Overall, these findings indicate that extreme heat increases the risk of mental-disorder related hospitalization in rural South Africa, both in the overall population and across key subgroups.

The short-term increase in admissions following extreme heat is consistent with evidence from Europe, North America, Asia, and Australia where high temperatures have been linked to increased mental health admissions.^2^ The proposed mechanisms span physiological, psychological, behavioral, and social pathways.^19^ Physiologically, heat exposure may disrupt thermoregulation, activate the hypothalamic-pituitary-adrenal axis and increase inflammation and oxidative stress.^19^ Psychologically, high temperatures have been associated with increased aggression, stress, heightened health anxiety, and reduced perceived control.^19^ Behaviorally, high temperature exposure has been linked with increased substance use, reduced adherence to therapeutic routines and sleep disruption.^19^ Socially, extreme heat can exacerbate isolation.^19^ Individuals with pre-existing mental disorders may be particularly vulnerable partly due to the effects of psychotropic medications on heat tolerance.^20^

Evidence on the relationship between heat and mental health in South Africa is limited. In a multi-country study, including South Africa, Kim *et al*. reported an increased suicide risk at higher ambient temperatures.^12^ Heat exposure has also been associated with increased violence and homicide rates in South Africa.^10,11^ Our findings extend this body of knowledge by providing the first direct estimates of the association between extreme heat and mental disorder-related hospital admissions in a rural South African setting. As our analysis is limited to a single hospital, generalizability is limited; however, the study provides observational evidence from a setting that is underrepresented in the literature. Multi-site and population-level studies are needed to determine whether these patterns hold across different regions in South Africa and the wider African context.

Our subgroup analysis indicated distinct responses by sex group to extreme heat. Females exhibited an increased risk of mental disorder admissions in the short term (lag 0-1), while males showed increased risk at delayed lags. We found no clear evidence in the literature of similar sex-specific lag patterns. Previous studies outside Africa have reported mixed findings, with some observing no sex differences^21^, and others suggesting greater vulnerability among men^22^ or women.^23^ These inconsistencies may reflect differences in study design (few studies examine long lags, potentially missing delayed responses), as well as variation in study populations, and climatic contexts. Behavioral and physiological factors, social roles, and the gendered division of labor have all been proposed as contributors to sex differences in heat-related wellbeing.^24^ Qualitative studies from South Africa indicate that traditional masculine norms, emphasizing stoicism, self-reliance, and emotional restraint, discourage health seeking from formal mental healthcare services and instead promote avoidance, substance use, or reliance on informal support such as friends and family.^25–27^ These patterns may help explain the delayed increase in admissions observed among men, as distress may accumulate before clinical presentation, whereas women may seek support earlier. Our findings indicate that females exhibited immediate, and males delayed heat-related admissions for mental disorders, reflecting potential physiological, behavioral, and social differences in vulnerability, that warrant further investigation.

We observed a delayed increase in mental disorder-related admissions among individuals aged <60 years and a decreased risk among those aged ≥60 years. Evidence from South Korea, the USA, and Vietnam indicated an increased risk of mental disorder-related hospital admissions or emergency department visits among older adults,^16,22,28^ potentially due to reduced thermoregulation and poorer physiological adaptation.^20^ However, Corvetto *et al*., reported a decreased risk of emergency department visits among Brazilians aged ≥65 years (cumulative lag 0-4) in response to extreme heat ^29^, suggesting that older adults may avoid seeking care during hot periods. In our study, the relatively small number of older adults with mental disorder-related admissions (156 from 1760 patients) warrants cautious interpretation. Increased risk of mental health admissions with elevated temperatures has been documented among younger and middle-aged individuals in China, Portugal, Switzerland, and the USA.^20,23,30,31^ Our findings suggest divergent heat-related risk patterns by age group, although they require cautious interpretation and confirmation in larger South African samples.

Given that schizophrenia accounted for 70.57% of all mental disorder admissions in Hlabisa, we stratified analyses by schizophrenia (ICD-10 F20-29) versus all other mental disorders (ICD-10 F00-19, F30-F99, X60-X84). We found no evidence that schizophrenia alone was associated with increased admissions in relation to extreme heat. This is unexpected, as studies from Australia, China, South Korea, and the USA have reported significant increases in hospitalizations and emergency department visits for schizophrenia during periods of high temperature.^20,21,28^ Psychotropic medications commonly prescribed for schizophrenia may partly explain these findings, as they can impair central thermoregulation and reduce heat tolerance.^20^ In Hlabisa, a rural setting in KwaZulu-Natal, individuals with schizophrenia may have lower medication coverage due to limited access to prescribing clinicians, transportation barriers, and reliance on informal or traditional care.^3^ These contextual factors may contribute to the absence of an observed association between heat and schizophrenia admissions. Further research is needed to determine whether similar patterns are observed in other rural regions of South Africa.

Our current study has several strengths, including the novel use of 13 years of mental health hospital admission data from a rural African setting and the application of distributed lag models to capture both immediate and delayed effects of extreme heat. Stratified analyses further provided insights into potentially vulnerable subgroups. Several limitations should be considered, including the single-center design, which may limit generalizability of findings to other contexts, small subgroup sample sizes, reliance on a single temperature metric, potential exposure misclassification, and the use of hospital admissions as a proxy for severe mental health outcomes, which may not reflect the full population burden.

This study provides novel evidence from rural South Africa demonstrating that extreme heat is associated with increased hospital admissions for mental disorders, with distinct temporal patterns by sex, age, and diagnosis. As South Africa is projected to experience further increases in the intensity, duration, and frequency of extreme heat, these results underscore the need to integrate mental health support into climate adaptation and public health strategies.^13^ Expanded national and multi-site studies are needed to confirm the present findings, better characterize population-level risks, and inform targeted interventions for vulnerable groups.

## Data Availability

Data on hospital admissions are available on the Africa Health Research Institute data repository at this link: https://data.ahri.org/index.php/home. The data can be downloaded after completion of a data use agreement available on the repository.

## Acknowledgments

We thank the Department of Health and the staff at Hlabisa Hospital for their role in the generation of clinical records utilized in this study. Finally, we express our gratitude to the patients admitted to Hlabisa Hospital whose data were used for this research.

During the preparation of this work, the authors used large language model-based artificial intelligence tools (ChatGPT, Claude, Gemini) to improve language and readability, assist with code development, and support figure preparation. After using these tools, the authors reviewed and edited the content as necessary and take full responsibility for the content of the published article.

## Author contributions

AB and CI conceived the study. KH, TM, and DG were responsible for the implementation and maintenance of the Africa Health Research Institute (AHRI) Hospital Information System (HIS) at Hlabisa Hospital. AB, CI, PL, and EL contributed to the methodology for secondary analysis. A.B., G.B., V.C., C.I., A.K., A.S., F.T., and A.T.-G. secured funding and provided resources for the research. EL was responsible for data curation, extraction of hospital records, formal analysis, and visualization. PL and AB provided supervision throughout the study. PL provided foundational analysis scripts that EL adapted and extended, and guided EL in the analytical approach. EL wrote the first draft of the manuscript. All authors reviewed the original draft, provided scientific input and editing for the final draft, and accepted responsibility for the decision to submit for publication. EL, PL, and CI had access to the data, and all authors accessed and verified the data.

## Literature

1 Global, regional, and national burden of 12 mental disorders in 204 countries and territories, 1990–2019: a systematic analysis for the Global Burden of Disease Study 2019. Lancet Psychiatry 2022; 9: 137–50.

2 Thompson R, Lawrance EL, Roberts LF, et al. Ambient temperature and mental health: a systematic review and meta-analysis. Lancet Planet Health 2023; 7: e580–9.

3 Vergunst R. From global-to-local: rural mental health in South Africa. Glob Health Action 2018; 11: 1413916.

4 Kirkbride JB, Anglin DM, Colman I, et al. The social determinants of mental health and disorder: evidence, prevention and recommendations. World Psychiatry 2024; 23: 58–90.

5 Remien RH, Stirratt MJ, Nguyen N, Robbins RN, Pala AN, Mellins CA. Mental health and HIV/AIDS: the need for an integrated response. AIDS 2019; 33: 1411–20.

6 UNAIDS. South Africa 2024 - HIV and AIDS Estimates. UNAIDS. 2025; published online Aug 19. https://aidsinfo.unaids.org (accessed Aug 19, 2025).

7 World Bank. Country Profile - South Africa. World Bank, Poverty and Inequality Platform. 2025; published online Aug 19. https://pip.worldbank.org/country-profiles/ZAF (accessed Aug 19, 2025).

8 Herman AA, Stein DJ, Seedat S, et al. The South African Stress and Health (SASH) study: 12-month and lifetime prevalence of common mental disorders. South African Medical Journal 2008; 99.

9 Craig A, Rochat T, Naicker SN, et al. The prevalence of probable depression and probable anxiety, and associations with adverse childhood experiences and socio-demographics: A national survey in South Africa. Front Public Health 2022; 10. DOI:10.3389/fpubh.2022.986531.

10 Gates A, Klein M, Acquaotta F, Garland RM, Scovronick N. Short-term association between ambient temperature and homicide in South Africa: a case-crossover study. Environmental Health 2019; 18: 109.

11 Schutte FH, Breetzke GD. The influence of extreme weather conditions on the magnitude and spatial distribution of crime in Tshwane (2001–2006). South African Geographical Journal 2018; 100: 364–77.

12 Kim Y, Kim H, Gasparrini A, et al. Suicide and Ambient Temperature: A Multi-Country Multi-City Study. Environ Health Perspect 2019; 127: 117007.

13 McBride CM, Kruger AC, Johnston C, Dyson L. Projected changes in daily temperature extremes for selected locations over South Africa. Weather Clim Extrem 2025; 47. DOI:10.1016/j.wace.2025.100753.

14 Herbst K, Mngomezulu T, Gareta D. AHRI.HIS-Admissions. 2023.

15 Hersbach H, Bell B, Berrisford P, et al. ERA5 hourly data on single levels from 1940 to present. 2023. DOI:10.24381/cds.adbb2d47.

16 Yoo E hye, Eum Y, Gao Q, Chen K. Effect of extreme temperatures on daily emergency room visits for mental disorders. Environmental Science and Pollution Research 2021; 28: 39243–56.

17 Gasparrini A, Armstrong B, Kenward MG. Distributed lag non-linear models. Stat Med 2010; 29: 2224–34.

18 Gasparrini A. Distributed Lag Linear and Non-Linear Models in R: The Package dlnm. J Stat Softw 2011; 43: 1–20.

19 Baecker L, Iyengar U, Del Piccolo MC, Mechelli A. “Impacts of extreme heat on mental health: Systematic review and qualitative investigation of the underpinning mechanisms”. Journal of Climate Change and Health 2025; 22. DOI:10.1016/j.joclim.2025.100446.

20 Kirby N V., Tetzlaff EJ, Kidd SA, et al. Susceptibility of persons with schizophrenia to extreme heat: A critical review of physiological, behavioural, and social factors. Science of The Total Environment 2025; 995: 179965.

21 Yoo E, Eum Y, Roberts JE, Gao Q, Chen K. Association between extreme temperatures and emergency room visits related to mental disorders: A multi-region time-series study in New York, USA. Science of The Total Environment 2021; 792: 148246.

22 Trang PM, Rocklöv J, Giang KB, Kullgren G, Nilsson M. Heatwaves and Hospital Admissions for Mental Disorders in Northern Vietnam. PLoS One 2016; 11: e0155609.

23 Almendra R, Loureiro A, Silva G, Vasconcelos J, Santana P. Short-term impacts of air temperature on hospitalizations for mental disorders in Lisbon. Science of the Total Environment 2019; 647: 127–33.

24 Belloc I, Gimenez-Nadal JI, Molina JA. Extreme temperatures: Gender differences in well-being. J Behav Exp Econ 2025; 117. DOI:10.1016/j.socec.2025.102405.

25 Mogano NTH, Letsoalo DL, Oduaran CA. Effects of masculine culture on the mental health of Northern Sotho male youth. BMC Psychol 2025; 13: 605.

26 Masemola HC, Moodley S V., Shirinde J. Perceptions and attitudes of black men in a rural district of South Africa towards depression and its treatment. South African Family Practice 2022; 64: 5557.

27 Sithambaram V, Wagner C, Cassimjee N. Exploring South African Indian men’s understanding of depression. South African Journal of Psychiatry 2024; 30. DOI:10.4102/sajpsychiatry.v30i0.2300.

28 Lee S, Lee H, Myung W, Kim EJ, Kim H. Mental disease-related emergency admissions attributable to hot temperatures. Science of The Total Environment 2018; 616–617: 688–94.

29 Corvetto JF, Federspiel A, Sewe MO, Müller T, Bunker A, Sauerborn R. Impact of heat on mental health emergency visits: A time series study from all public emergency centres, in Curitiba, Brazil. BMJ Open 2023; 13. DOI:10.1136/bmjopen-2023-079049.

30 Niu L, Girma B, Liu B, Schinasi LH, Clougherty JE, Sheffield P. Temperature and mental health-related emergency department and hospital encounters among children, adolescents and young adults. Epidemiol Psychiatr Sci 2023; 32. DOI:10.1017/S2045796023000161.

31 Bundo M, de Schrijver E, Federspiel A, et al. Ambient temperature and mental health hospitalizations in Bern, Switzerland: A 45-year time-series study. PLoS One 2021; 16. DOI:10.1371/journal.pone.0258302.

