## SupplementaryInformation for "The Effects of Extreme Heat on Mental Disorder Admissions in Hlabisa, South Africa: A 13-Year Time-Series Analysis"

**Supplementary Information**

**List of Supplementary Tables**

**Table S1.** Top 10 model configurations for ERA5 data

**Table S2.** Top 10 model configurations for Riverview weather station data

**Table S3.** Top 10 model configurations from extended grid search including seasonality degrees of freedom

**Table S4.** Cumulative relative risk of hospital admission for 95th, 97.5th, and 99th percentile of daily maximum temperature for ERA5 and weather station data

**Table S5.** Cumulative relative risk of hospital admission for 95th, 97.5th, and 99th percentile of daily maximum temperature for stratified data (Sex, Age)

**Table S6.** Cumulative relative risk of hospital admission for 95th, 97.5th, and 99th percentile of daily maximum temperature for stratified data (Schizophrenia & Other mental health)

**Table S7.** Cumulative relative risk of hospital admission for 95th, 97.5th, and 99th percentile of daily maximum temperature for ERA5 data (sensitivity analysis - seasonality degrees of freedom optimized)

**Table S8.** Lag-specific relative risk of hospital admission for 95th, 97.5th, and 99th percentile of daily maximum temperature for ERA5 and weather station data

**Table S9.** Lag-specific relative risk of hospital admission for 95th, 97.5th, and 99th percentile of daily maximum temperature for ERA5 stratified data

**Table S10.** Lag-specific temperature effects (Schizophrenia & Other mental health)

**Table S11.** Lag-specific temperature effects (sensitivity analysis - seasonality degrees of freedom optimized)

**List of Supplementary Figures**

**Figure S1.** Cumulative exposure-response curves of maximum daily temperature (°C), based on ERA5 data, and cumulative relative risk of admission for mental disorder (relative to the median temperature)

**Figure S2.** Cumulative (left) and lag-specific (right) exposure-response curves for maximum daily temperature (°C) and relative risk of admission for mental disorder (relative to the median temperature) for all ERA5 data

**Figure S3.** Cumulative (left) and lag-specific (right) exposure-response curves for maximum daily temperature (°C) and relative risk of admission for mental disorder (relative to the median temperature) for all Riverview station data

**Figure S4.** Cumulative (left) and lag-specific (right) exposure-response curves for maximum daily temperature (°C) and relative risk of admission for mental disorder (relative to the median temperature) for females

**Figure S5.** Cumulative (left) and lag-specific (right) exposure-response curves for maximum daily temperature (°C) and relative risk of admission for mental disorder (relative to the median temperature) for males

**Figure S6.** Cumulative (left) and lag-specific (right) exposure-response curves for maximum daily temperature (°C) and relative risk of admission for mental disorder (relative to the median temperature) for age <60

**Figure S7.** Cumulative (left) and lag-specific (right) exposure-response curves for maximum daily temperature (°C) and relative risk of admission for mental disorder (relative to the median temperature) for individuals age ≥60

**Figure S8.** Cumulative (left) and lag-specific (right) exposure-response curves for maximum daily temperature (°C) and relative risk of admission for mental disorder (relative to the median temperature) for individuals diagnosed with schizophrenia

**Figure S9.** Cumulative (left) and lag-specific (right) exposure-response curves for maximum daily temperature (°C) and relative risk of admission for mental disorder (relative to the median temperature) for individuals diagnosed with mental health diagnoses other than schizophrenia

**Figure S10.** Cumulative (left) and lag-specific (right) exposure-response curves for maximum daily temperature (°C) and relative risk of admission for mental disorder (relative to the median temperature) for all ERA5 data (sensitivity analysis - seasonality degrees of freedom optimized)

### Table S1. Top 10 model configurations for ERA5 data

| **Temp knot 1** | **Temp knot 2** | **Lag knot 1** | **Lag knot 2** | **QAIC** | **AIC** | **Overdispersion** |
| --- | --- | --- | --- | --- | --- | --- |
| 30 | 33 | 14 |  | 7277.59 | 7254.44 | 1.11 |
| 29 | 33 | 14 |  | 7277.68 | 7254.65 | 1.11 |
| 30 | 32 | 14 |  | 7277.71 | 7254.68 | 1.11 |
| 28 | 33 | 14 |  | 7277.77 | 7254.85 | 1.11 |
| 29 | 32 | 14 |  | 7277.79 | 7254.85 | 1.11 |
| 30 | 31 | 14 |  | 7277.79 | 7254.85 | 1.11 |
| 27 | 33 | 14 |  | 7277.85 | 7255.02 | 1.11 |
| 30 | 33 | 7 | 14 | 7277.87 | 7254.77 | 1.10 |
| 28 | 32 | 14 |  | 7277.87 | 7255.02 | 1.11 |
| 29 | 31 | 14 |  | 7277.88 | 7255.01 | 1.11 |

### Table S2. Top 10 model configurations for Riverview weather station data

| **Temp knot 1** | **Temp knot 2** | **Lag knot 1** | **Lag knot 2** | **QAIC** | **AIC** | **Overdispersion** |
| --- | --- | --- | --- | --- | --- | --- |
| 30 | 33 | 14 |  | 7271.54 | 7251.33 | 1.09 |
| 30 | 32 | 14 |  | 7271.62 | 7251.42 | 1.09 |
| 29 | 33 | 14 |  | 7271.83 | 7251.56 | 1.09 |
| 30 | 31 | 14 |  | 7271.85 | 7251.59 | 1.09 |
| 29 | 32 | 14 |  | 7272.02 | 7251.71 | 1.09 |
| 28 | 33 | 14 |  | 7272.15 | 7251.78 | 1.10 |
| 27 | 33 | 14 |  | 7272.35 | 7251.90 | 1.10 |
| 29 | 31 | 14 |  | 7272.37 | 7251.94 | 1.10 |
| 28 | 32 | 14 |  | 7272.40 | 7251.95 | 1.10 |
| 26 | 33 | 14 |  | 7272.40 | 7251.91 | 1.10 |

Table S3. Top 10 model configurations from extended grid search including seasonality degrees of freedom.

| Seasonality df | Temp knot 1 | Temp knot 2 | Lag  knot 1 | Lag  knot 2 | QAIC | AIC | Overdispersion |
| --- | --- | --- | --- | --- | --- | --- | --- |
| 49 | 26 | 27 | 14 |  | 7244.50 | 7235.63 | 1.07 |
| 49 | 26 | 28 | 14 |  | 7244.83 | 7235.93 | 1.07 |
| 49 | 27 | 28 | 14 |  | 7245.16 | 7236.22 | 1.07 |
| 49 | 26 | 29 | 14 |  | 7245.17 | 7236.23 | 1.07 |
| 49 | 26 | 27 | 6 |  | 7245.43 | 7236.58 | 1.07 |
| 49 | 26 | 30 | 14 |  | 7245.47 | 7236.49 | 1.07 |
| 49 | 27 | 29 | 14 |  | 7245.49 | 7236.51 | 1.07 |
| 49 | 26 | 31 | 14 |  | 7245.72 | 7236.69 | 1.07 |
| 49 | 26 | 28 | 6 |  | 7245.75 | 7236.87 | 1.07 |
| 49 | 27 | 30 | 14 |  | 7245.78 | 7236.75 | 1.07 |

### Table S4. Cumulative relative risk of hospital admission for 95^th^, 97.5^th^, and 99^th^ percentile of daily maximum temperature for ERA5 and weather station data

| **Lag** | **All – ERA5** | | | **All – Riverview WS** | | |
| --- | --- | --- | --- | --- | --- | --- |
|  | **95th** | **97.5th** | **99th** | **95th** | **97.5th** | **99th** |
| 0-3 | 1.24 (0.98–1.57) | 1.35 (1–1.81) | 1.47 (0.93–2.32) | 1.21 (0.93–1.58) | 1.35 (0.99–1.82) | 1.64 (0.99–2.7) |
| 0-7 | 1.31 (0.91–1.89) | 1.46 (0.93–2.31) | 1.65 (0.81–3.36) | 1.24 (0.82–1.87) | 1.44 (0.9–2.3) | 1.94 (0.89–4.21) |
| 0-14 | 1.15 (0.68–1.96) | 1.29 (0.66–2.52) | 1.49 (0.51–4.37) | 1.02 (0.55–1.9) | 1.22 (0.61–2.44) | 1.82 (0.57–5.83) |
| 0-21 | 1.13 (0.56–2.28) | 1.52 (0.64–3.6) | 2.21 (0.55–8.91) | 0.98 (0.43–2.24) | 1.41 (0.57–3.48) | 3.13 (0.71–13.72) |

### Table S5. Cumulative relative risk of hospital admission for 95^th^, 97.5^th^, and 99^th^ percentile of daily maximum temperature for stratified data (Sex, Age)

| **Lag** | **Female** | | | **Male** | | | **Age <60** | | | **Age ≥60** | | |
| --- | --- | --- | --- | --- | --- | --- | --- | --- | --- | --- | --- | --- |
|  | **95th** | **97.5th** | **99th** | **95th** | **97.5th** | **99th** | **95th** | **97.5th** | **99th** | **95th** | **97.5th** | **99th** |
| 0-3 | 1.44 (1.02–2.04)* | 1.49 (0.96–2.31) | 1.5 (0.75–2.98) | 1.06 (0.77–1.47) | 1.2 (0.81–1.79) | 1.41 (0.77–2.56) | 1.18 (0.92–1.52) | 1.34 (0.98–1.82) | 1.55 (0.97–2.49) | 1.54 (0.75–3.16) | 0.93 (0.34–2.56) | 0.45 (0.08–2.42) |
| 0-7 | 1.47 (0.86–2.52) | 1.62 (0.82–3.18) | 1.76 (0.6–5.13) | 1.13 (0.69–1.85) | 1.29 (0.7–2.36) | 1.5 (0.59–3.84) | 1.24 (0.84–1.81) | 1.47 (0.92–2.36) | 1.82 (0.87–3.8) | 1.47 (0.48–4.45) | 0.66 (0.14–3.14) | 0.22 (0.02–3.01) |
| 0-14 | 0.92 (0.41–2.03) | 1.15 (0.42–3.12) | 1.56 (0.31–7.87) | 1.31 (0.64–2.67) | 1.35 (0.55–3.28) | 1.37 (0.33–5.73) | 1.14 (0.65–1.98) | 1.39 (0.69–2.79) | 1.81 (0.59–5.51) | 0.58 (0.11–3) | 0.2 (0.02–1.86) | 0.05 (0–2.35) |
| 0-21 | 0.64 (0.22–1.82) | 0.85 (0.23–3.19) | 1.32 (0.16–11.12) | 1.7 (0.67–4.31) | 2.25 (0.72–7.04) | 3.09 (0.5–19.31) | 1.2 (0.58–2.49) | 1.86 (0.76–4.59) | 3.28 (0.77–13.95) | 0.18 (0.02–1.58) | 0.03 (0–0.58)† | 0 (0–0.56)† |

Note: *Significant Increase; †Significant decrease

### Table S6. Cumulative relative risk of hospital admission for 95th, 97.5th, and 99th percentile of daily maximum temperature for stratified data (Schizophrenia & Other mental health)

| **Lag** | **Schizophrenia** | | | **Other Mental Health** | | |
| --- | --- | --- | --- | --- | --- | --- |
|  | **95th** | **97.5th** | **99th** | **95th** | **97.5th** | **99th** |
| 0-3 | 0.99 (0.75–1.32) | 1.15 (0.81–1.63) | 1.39 (0.82–2.36) | 1.99 (1.32–2.99)* | 1.79 (1.06–3.02)* | 1.45 (0.63–3.34) |
| 0-7 | 0.93 (0.6–1.43) | 1.14 (0.67–1.95) | 1.51 (0.66–3.45) | 2.77 (1.48–5.17)* | 2.27 (1.02–5.06)* | 1.58 (0.43–5.76) |
| 0-14 | 0.78 (0.41–1.47) | 0.99 (0.45–2.18) | 1.41 (0.4–4.92) | 2.76 (1.11–6.85)* | 1.98 (0.62–6.38) | 1.15 (0.16–8.18) |
| 0-21 | 0.8 (0.35–1.84) | 1.31 (0.47–3.64) | 2.57 (0.51–13.01) | 2.64 (0.8–8.73) | 1.72 (0.38–7.78) | 0.88 (0.07–11.21) |

Note: *Significant Increase; †Significant decrease

### Table S7. Cumulative relative risk of hospital admission for 95^th^, 97.5^th^, and 99^th^ percentile of daily maximum temperature for ERA5 data (sensitivity analysis - seasonality degrees of freedom optimized).

| **Lag** | **95th** | **97.5th** | **99th** |
| --- | --- | --- | --- |
| 0-3 | 1.23 (1–1.51)* | 1.3 (0.97–1.75) | 1.37 (0.92–2.05) |
| 0-7 | 1.27 (0.95–1.72) | 1.32 (0.86–2.05) | 1.37 (0.75–2.49) |
| 0-14 | 1.09 (0.72–1.64) | 1 (0.54–1.85) | 0.91 (0.39–2.13) |
| 0-21 | 1.15 (0.69–1.9) | 1.05 (0.49–2.25) | 0.94 (0.32–2.73) |

Note: *Significant Increase; †Significant decrease

### Table S8. Lag-specific relative risk of hospital admission for 95^th^, 97.5^th^, and 99^th^ percentile of daily maximum temperature for ERA5 and weather station data

| **Lag** | **All – ERA5** | | | **All – Riverview WS** | | |
| --- | --- | --- | --- | --- | --- | --- |
|  | **95th** | **97.5th** | **99th** | **95th** | **97.5th** | **99th** |
| 0 | 1.07 (0.99–1.15) | 1.1 (1–1.2)* | 1.13 (0.99–1.29) | 1.07 (0.99–1.16) | 1.11 (1.01–1.22)* | 1.18 (1–1.39)* |
| 1 | 1.06 (0.99–1.13) | 1.08 (1–1.17)* | 1.11 (0.99–1.24) | 1.06 (0.98–1.13) | 1.09 (1–1.19)* | 1.15 (1–1.33) |
| 2 | 1.05 (0.99–1.11) | 1.07 (1–1.14) | 1.09 (0.98–1.2) | 1.04 (0.98–1.11) | 1.07 (1–1.15) | 1.13 (0.99–1.27) |
| 3 | 1.04 (0.99–1.09) | 1.05 (0.99–1.11) | 1.07 (0.98–1.17) | 1.03 (0.98–1.09) | 1.06 (0.99–1.13) | 1.1 (0.99–1.23) |
| 4 | 1.03 (0.98–1.07) | 1.04 (0.99–1.09) | 1.05 (0.97–1.14) | 1.02 (0.97–1.07) | 1.04 (0.98–1.1) | 1.08 (0.98–1.19) |
| 5 | 1.02 (0.98–1.06) | 1.03 (0.98–1.07) | 1.03 (0.96–1.11) | 1.01 (0.97–1.06) | 1.03 (0.97–1.08) | 1.06 (0.96–1.16) |
| 6 | 1.01 (0.97–1.05) | 1.01 (0.97–1.06) | 1.02 (0.95–1.1) | 1 (0.96–1.05) | 1.01 (0.96–1.07) | 1.04 (0.95–1.14) |
| 7 | 1 (0.96–1.04) | 1 (0.96–1.05) | 1.01 (0.93–1.09) | 0.99 (0.95–1.04) | 1 (0.95–1.05) | 1.02 (0.93–1.12) |
| 8 | 0.99 (0.95–1.03) | 0.99 (0.95–1.04) | 1 (0.92–1.08) | 0.98 (0.94–1.03) | 0.99 (0.94–1.05) | 1.01 (0.91–1.11) |
| 9 | 0.99 (0.95–1.03) | 0.99 (0.94–1.04) | 0.99 (0.91–1.07) | 0.98 (0.93–1.03) | 0.98 (0.93–1.04) | 1 (0.9–1.1) |
| 10 | 0.98 (0.94–1.03) | 0.98 (0.93–1.03) | 0.98 (0.9–1.07) | 0.97 (0.93–1.02) | 0.98 (0.92–1.04) | 0.99 (0.89–1.1) |
| 11 | 0.98 (0.94–1.02) | 0.98 (0.93–1.03) | 0.98 (0.9–1.07) | 0.97 (0.92–1.02) | 0.97 (0.92–1.03) | 0.99 (0.89–1.1) |
| 12 | 0.98 (0.93–1.02) | 0.98 (0.93–1.03) | 0.98 (0.9–1.07) | 0.97 (0.92–1.02) | 0.97 (0.91–1.03) | 0.99 (0.89–1.1) |
| 13 | 0.98 (0.93–1.02) | 0.98 (0.93–1.03) | 0.98 (0.9–1.07) | 0.97 (0.92–1.02) | 0.97 (0.92–1.03) | 0.99 (0.89–1.1) |
| 14 | 0.98 (0.94–1.02) | 0.98 (0.93–1.03) | 0.99 (0.91–1.07) | 0.97 (0.92–1.02) | 0.98 (0.92–1.03) | 1 (0.91–1.1) |
| 15 | 0.98 (0.94–1.02) | 0.99 (0.94–1.03) | 1 (0.93–1.08) | 0.97 (0.93–1.02) | 0.99 (0.93–1.04) | 1.01 (0.92–1.11) |
| 16 | 0.98 (0.94–1.02) | 0.99 (0.95–1.04) | 1.01 (0.94–1.09) | 0.98 (0.93–1.03) | 1 (0.94–1.05) | 1.03 (0.94–1.13) |
| 17 | 0.99 (0.95–1.03) | 1 (0.96–1.05) | 1.03 (0.95–1.11) | 0.99 (0.94–1.04) | 1.01 (0.95–1.07) | 1.06 (0.96–1.16) |
| 18 | 0.99 (0.95–1.04) | 1.02 (0.96–1.07) | 1.05 (0.96–1.15) | 0.99 (0.94–1.05) | 1.02 (0.96–1.09) | 1.08 (0.98–1.2) |
| 19 | 1 (0.94–1.06) | 1.03 (0.96–1.1) | 1.07 (0.97–1.19) | 1 (0.94–1.07) | 1.04 (0.96–1.12) | 1.11 (0.99–1.26) |
| 20 | 1.01 (0.94–1.08) | 1.04 (0.96–1.13) | 1.1 (0.97–1.24) | 1.01 (0.94–1.09) | 1.06 (0.97–1.15) | 1.15 (0.99–1.33) |
| 21 | 1.01 (0.94–1.1) | 1.06 (0.96–1.16) | 1.12 (0.97–1.3) | 1.02 (0.94–1.12) | 1.08 (0.97–1.19) | 1.18 (0.99–1.4) |

Note: *Significant Increase; †Significant decrease

### Table S9. Lag-specific relative risk of hospital admission for 95^th^, 97.5^th^, and 99^th^ percentile of daily maximum temperature for ERA5 stratified data

| **Lag** | **Female** | | | **Male** | | | **Age <60** | | | **Age ≥60** | | |
| --- | --- | --- | --- | --- | --- | --- | --- | --- | --- | --- | --- | --- |
|  | **95th** | **97.5th** | **99th** | **95th** | **97.5th** | **99th** | **95th** | **97.5th** | **99th** | **95th** | **97.5th** | **99th** |
| 0 | 1.13 (1.02–1.26)* | 1.14 (1–1.3)* | 1.13 (0.93–1.39) | 1.01 (0.92–1.12) | 1.05 (0.94–1.18) | 1.11 (0.94–1.33) | 1.05 (0.97–1.13) | 1.09 (1–1.19) | 1.14 (1–1.31) | 1.18 (0.96–1.47) | 1.04 (0.78–1.38) | 0.83 (0.51–1.35) |
| 1 | 1.11 (1.01–1.21)* | 1.12 (1–1.25) | 1.12 (0.94–1.33) | 1.01 (0.93–1.1) | 1.05 (0.94–1.16) | 1.09 (0.94–1.27) | 1.04 (0.98–1.11) | 1.08 (1–1.16) | 1.12 (0.99–1.26) | 1.15 (0.95–1.38) | 1.02 (0.79–1.31) | 0.84 (0.55–1.28) |
| 2 | 1.08 (1–1.17) | 1.09 (0.99–1.2) | 1.1 (0.94–1.28) | 1.01 (0.94–1.09) | 1.04 (0.95–1.13) | 1.07 (0.94–1.23) | 1.03 (0.98–1.1) | 1.06 (0.99–1.14) | 1.1 (0.99–1.22) | 1.11 (0.94–1.31) | 1 (0.8–1.24) | 0.84 (0.58–1.22) |
| 3 | 1.06 (0.99–1.13) | 1.07 (0.98–1.16) | 1.08 (0.94–1.23) | 1.01 (0.95–1.08) | 1.03 (0.96–1.11) | 1.06 (0.94–1.19) | 1.03 (0.98–1.08) | 1.05 (0.99–1.11) | 1.08 (0.99–1.18) | 1.07 (0.93–1.24) | 0.98 (0.81–1.18) | 0.84 (0.61–1.17) |
| 4 | 1.03 (0.97–1.1) | 1.05 (0.97–1.13) | 1.06 (0.94–1.2) | 1.01 (0.96–1.07) | 1.02 (0.96–1.1) | 1.04 (0.93–1.15) | 1.02 (0.98–1.07) | 1.04 (0.98–1.09) | 1.06 (0.98–1.15) | 1.04 (0.91–1.18) | 0.96 (0.81–1.13) | 0.84 (0.63–1.13) |
| 5 | 1.01 (0.96–1.07) | 1.03 (0.96–1.1) | 1.05 (0.93–1.17) | 1.02 (0.96–1.07) | 1.02 (0.96–1.08) | 1.02 (0.93–1.13) | 1.01 (0.97–1.06) | 1.03 (0.98–1.08) | 1.05 (0.97–1.13) | 1.01 (0.89–1.14) | 0.94 (0.81–1.1) | 0.85 (0.65–1.11) |
| 6 | 0.99 (0.94–1.05) | 1.01 (0.94–1.08) | 1.03 (0.92–1.15) | 1.02 (0.97–1.07) | 1.01 (0.95–1.08) | 1.01 (0.92–1.11) | 1.01 (0.97–1.05) | 1.02 (0.97–1.07) | 1.03 (0.95–1.11) | 0.98 (0.87–1.1) | 0.92 (0.79–1.07) | 0.85 (0.65–1.09) |
| 7 | 0.98 (0.92–1.03) | 0.99 (0.93–1.06) | 1.02 (0.91–1.14) | 1.02 (0.97–1.07) | 1.01 (0.95–1.07) | 1 (0.9–1.1) | 1 (0.96–1.04) | 1.01 (0.96–1.06) | 1.02 (0.94–1.1) | 0.95 (0.85–1.07) | 0.91 (0.78–1.05) | 0.84 (0.65–1.09) |
| 8 | 0.96 (0.9–1.02) | 0.98 (0.91–1.05) | 1 (0.89–1.13) | 1.02 (0.96–1.07) | 1.01 (0.94–1.07) | 0.99 (0.89–1.1) | 1 (0.95–1.04) | 1 (0.95–1.05) | 1.01 (0.93–1.09) | 0.93 (0.82–1.05) | 0.89 (0.76–1.04) | 0.84 (0.64–1.1) |
| 9 | 0.95 (0.89–1.01) | 0.96 (0.89–1.04) | 0.99 (0.88–1.12) | 1.02 (0.96–1.08) | 1.01 (0.94–1.08) | 0.98 (0.88–1.1) | 0.99 (0.95–1.04) | 0.99 (0.94–1.05) | 1 (0.92–1.09) | 0.91 (0.8–1.03) | 0.87 (0.74–1.03) | 0.84 (0.63–1.1) |
| 10 | 0.94 (0.88–1)† | 0.95 (0.88–1.03) | 0.98 (0.87–1.12) | 1.02 (0.96–1.08) | 1 (0.94–1.08) | 0.98 (0.88–1.1) | 0.99 (0.94–1.03) | 0.99 (0.94–1.05) | 0.99 (0.91–1.09) | 0.89 (0.78–1.02) | 0.86 (0.73–1.01) | 0.83 (0.62–1.1) |
| 11 | 0.93 (0.87–0.99)† | 0.94 (0.87–1.02) | 0.98 (0.86–1.11) | 1.02 (0.96–1.08) | 1.01 (0.94–1.08) | 0.98 (0.88–1.1) | 0.99 (0.94–1.03) | 0.99 (0.93–1.04) | 0.99 (0.91–1.08) | 0.88 (0.76–1) | 0.85 (0.71–1) | 0.82 (0.61–1.09) |
| 12 | 0.92 (0.86–0.99)† | 0.94 (0.87–1.02) | 0.97 (0.85–1.1) | 1.02 (0.96–1.09) | 1.01 (0.94–1.08) | 0.99 (0.88–1.11) | 0.98 (0.94–1.03) | 0.99 (0.93–1.04) | 0.99 (0.91–1.09) | 0.86 (0.75–0.99)† | 0.83 (0.7–0.99)† | 0.81 (0.6–1.08) |
| 13 | 0.92 (0.86–0.98)† | 0.93 (0.86–1.01) | 0.97 (0.85–1.1) | 1.02 (0.97–1.09) | 1.01 (0.95–1.09) | 1 (0.89–1.11) | 0.98 (0.94–1.03) | 0.99 (0.94–1.04) | 1 (0.91–1.09) | 0.85 (0.75–0.98)† | 0.82 (0.7–0.97)† | 0.79 (0.6–1.05) |
| 14 | 0.92 (0.86–0.98)† | 0.93 (0.87–1) | 0.96 (0.85–1.09) | 1.03 (0.97–1.09) | 1.02 (0.96–1.09) | 1.01 (0.91–1.12) | 0.99 (0.94–1.03) | 0.99 (0.94–1.05) | 1.01 (0.92–1.09) | 0.85 (0.74–0.96)† | 0.81 (0.69–0.95)† | 0.78 (0.59–1.02) |
| 15 | 0.92 (0.87–0.98)† | 0.93 (0.87–1) | 0.96 (0.86–1.08) | 1.03 (0.97–1.08) | 1.03 (0.97–1.1) | 1.03 (0.93–1.14) | 0.99 (0.95–1.03) | 1 (0.95–1.05) | 1.02 (0.94–1.1) | 0.84 (0.75–0.96)† | 0.8 (0.69–0.93)† | 0.76 (0.58–0.99)† |
| 16 | 0.93 (0.87–0.99)† | 0.94 (0.87–1.01) | 0.96 (0.86–1.08) | 1.03 (0.98–1.09) | 1.04 (0.98–1.11) | 1.05 (0.95–1.16) | 0.99 (0.95–1.03) | 1.01 (0.96–1.06) | 1.04 (0.96–1.12) | 0.84 (0.75–0.96)† | 0.79 (0.68–0.92)† | 0.74 (0.56–0.96)† |
| 17 | 0.94 (0.88–1)† | 0.95 (0.88–1.02) | 0.97 (0.86–1.09) | 1.03 (0.98–1.09) | 1.05 (0.99–1.12) | 1.08 (0.98–1.19) | 1 (0.95–1.04) | 1.02 (0.97–1.07) | 1.06 (0.97–1.14) | 0.85 (0.74–0.97)† | 0.78 (0.66–0.93)† | 0.71 (0.54–0.95)† |
| 18 | 0.95 (0.88–1.02) | 0.95 (0.88–1.04) | 0.97 (0.85–1.12) | 1.03 (0.97–1.1) | 1.07 (0.99–1.15) | 1.11 (1–1.24) | 1 (0.95–1.05) | 1.03 (0.98–1.1) | 1.08 (0.99–1.18) | 0.85 (0.73–0.99)† | 0.77 (0.64–0.94)† | 0.69 (0.5–0.96)† |
| 19 | 0.96 (0.88–1.04) | 0.96 (0.87–1.07) | 0.98 (0.83–1.15) | 1.04 (0.96–1.12) | 1.08 (0.99–1.18) | 1.15 (1.01–1.31)* | 1.01 (0.95–1.07) | 1.05 (0.98–1.12) | 1.11 (0.99–1.23) | 0.86 (0.72–1.02) | 0.77 (0.61–0.96)† | 0.67 (0.45–0.98)† |
| 20 | 0.97 (0.88–1.07) | 0.97 (0.86–1.1) | 0.98 (0.81–1.19) | 1.04 (0.95–1.14) | 1.1 (0.99–1.22) | 1.19 (1.02–1.38)* | 1.02 (0.95–1.09) | 1.06 (0.98–1.16) | 1.13 (1–1.29) | 0.87 (0.7–1.06) | 0.76 (0.58–1)† | 0.64 (0.41–1.02) |
| 21 | 0.98 (0.87–1.11) | 0.99 (0.85–1.14) | 0.99 (0.79–1.24) | 1.04 (0.93–1.16) | 1.12 (0.98–1.26) | 1.23 (1.02–1.47)* | 1.02 (0.94–1.11) | 1.08 (0.98–1.19) | 1.16 (1–1.35)* | 0.87 (0.68–1.11) | 0.75 (0.55–1.04) | 0.62 (0.36–1.06) |

Note: *Significant Increase; †Significant decreas

### Table S10. Lag-specific temperature effects (Schizophrenia & Other mental health)

| **Lag** | **Schizophrenia** | | | **Other Mental Health** | | |
| --- | --- | --- | --- | --- | --- | --- |
|  | **95th** | **97.5th** | **99th** | **95th** | **97.5th** | **99th** |
| 0 | 1 (0.92–1.09) | 1.04 (0.94–1.15) | 1.11 (0.95–1.29) | 1.23 (1.09–1.4)* | 1.21 (1.04–1.4)* | 1.14 (0.89–1.45) |
| 1 | 1 (0.92–1.07) | 1.03 (0.94–1.13) | 1.09 (0.95–1.25) | 1.2 (1.08–1.34)* | 1.18 (1.03–1.34)* | 1.12 (0.9–1.38) |
| 2 | 0.99 (0.93–1.06) | 1.02 (0.95–1.11) | 1.07 (0.95–1.2) | 1.17 (1.07–1.29)* | 1.15 (1.03–1.29)* | 1.09 (0.91–1.32) |
| 3 | 0.99 (0.93–1.05) | 1.01 (0.95–1.08) | 1.05 (0.95–1.17) | 1.15 (1.06–1.25)* | 1.12 (1.02–1.24)* | 1.07 (0.91–1.26) |
| 4 | 0.99 (0.94–1.04) | 1.01 (0.95–1.07) | 1.04 (0.95–1.14) | 1.12 (1.04–1.21)* | 1.1 (1.01–1.2)* | 1.05 (0.91–1.22) |
| 5 | 0.98 (0.94–1.03) | 1 (0.94–1.05) | 1.02 (0.94–1.12) | 1.1 (1.03–1.17)* | 1.08 (0.99–1.17) | 1.03 (0.9–1.18) |
| 6 | 0.98 (0.94–1.03) | 0.99 (0.94–1.05) | 1.01 (0.93–1.1) | 1.08 (1.01–1.15)* | 1.06 (0.98–1.14) | 1.02 (0.89–1.16) |
| 7 | 0.98 (0.93–1.02) | 0.99 (0.93–1.04) | 1 (0.92–1.09) | 1.06 (0.99–1.13) | 1.04 (0.96–1.12) | 1 (0.87–1.15) |
| 8 | 0.98 (0.93–1.02) | 0.98 (0.93–1.04) | 0.99 (0.91–1.09) | 1.04 (0.97–1.11) | 1.02 (0.94–1.11) | 0.99 (0.86–1.14) |
| 9 | 0.97 (0.93–1.02) | 0.98 (0.92–1.04) | 0.99 (0.9–1.08) | 1.02 (0.95–1.1) | 1 (0.92–1.1) | 0.97 (0.84–1.13) |
| 10 | 0.97 (0.92–1.03) | 0.98 (0.92–1.04) | 0.98 (0.89–1.08) | 1.01 (0.93–1.09) | 0.99 (0.9–1.08) | 0.96 (0.83–1.12) |
| 11 | 0.97 (0.92–1.03) | 0.98 (0.92–1.04) | 0.98 (0.89–1.09) | 1 (0.92–1.08) | 0.98 (0.89–1.08) | 0.96 (0.82–1.12) |
| 12 | 0.97 (0.92–1.03) | 0.98 (0.92–1.04) | 0.98 (0.89–1.09) | 0.99 (0.91–1.07) | 0.97 (0.88–1.07) | 0.95 (0.81–1.11) |
| 13 | 0.98 (0.93–1.03) | 0.98 (0.92–1.04) | 0.99 (0.9–1.09) | 0.98 (0.91–1.06) | 0.97 (0.88–1.06) | 0.94 (0.81–1.1) |
| 14 | 0.98 (0.93–1.03) | 0.99 (0.93–1.05) | 1 (0.91–1.1) | 0.98 (0.91–1.05) | 0.96 (0.88–1.05) | 0.94 (0.81–1.09) |
| 15 | 0.98 (0.94–1.03) | 1 (0.94–1.05) | 1.02 (0.93–1.11) | 0.98 (0.91–1.05) | 0.96 (0.89–1.05) | 0.95 (0.82–1.09) |
| 16 | 0.99 (0.94–1.04) | 1.01 (0.95–1.06) | 1.03 (0.95–1.13) | 0.98 (0.91–1.05) | 0.97 (0.89–1.05) | 0.95 (0.83–1.09) |
| 17 | 0.99 (0.94–1.04) | 1.02 (0.96–1.08) | 1.05 (0.96–1.15) | 0.99 (0.92–1.06) | 0.97 (0.89–1.06) | 0.95 (0.83–1.1) |
| 18 | 1 (0.94–1.06) | 1.03 (0.97–1.1) | 1.08 (0.98–1.19) | 0.99 (0.91–1.08) | 0.98 (0.89–1.08) | 0.96 (0.82–1.13) |
| 19 | 1.01 (0.94–1.07) | 1.05 (0.97–1.13) | 1.11 (0.98–1.24) | 1 (0.91–1.11) | 0.99 (0.88–1.11) | 0.97 (0.8–1.18) |
| 20 | 1.01 (0.94–1.1) | 1.06 (0.97–1.17) | 1.14 (0.99–1.31) | 1.01 (0.9–1.14) | 1 (0.86–1.15) | 0.98 (0.78–1.23) |
| 21 | 1.02 (0.93–1.12) | 1.08 (0.97–1.21) | 1.17 (0.99–1.38) | 1.02 (0.88–1.17) | 1.01 (0.85–1.2) | 0.99 (0.75–1.3) |

Note: *Significant Increase; †Significant decrease

### Table S11. Lag-specific temperature effects (sensitivity analysis - seasonality degrees of freedom optimized).

| **Lag** | **95th** | **97.5th** | **99th** |
| --- | --- | --- | --- |
| 0 | 1.07 (1.01–1.14)* | 1.09 (1–1.19) | 1.12 (0.99–1.26) |
| 1 | 1.06 (1–1.11)* | 1.07 (1–1.16) | 1.09 (0.98–1.21) |
| 2 | 1.04 (1–1.09) | 1.06 (0.99–1.13) | 1.07 (0.98–1.17) |
| 3 | 1.03 (1–1.07) | 1.04 (0.99–1.1) | 1.05 (0.97–1.13) |
| 4 | 1.02 (0.99–1.06) | 1.03 (0.98–1.08) | 1.03 (0.96–1.1) |
| 5 | 1.01 (0.98–1.04) | 1.01 (0.97–1.06) | 1.01 (0.95–1.07) |
| 6 | 1 (0.98–1.03) | 1 (0.96–1.04) | 0.99 (0.93–1.05) |
| 7 | 1 (0.97–1.03) | 0.99 (0.94–1.03) | 0.98 (0.92–1.04) |
| 8 | 0.99 (0.96–1.02) | 0.98 (0.93–1.02) | 0.96 (0.9–1.03) |
| 9 | 0.98 (0.95–1.02) | 0.97 (0.92–1.02) | 0.95 (0.89–1.02) |
| 10 | 0.98 (0.95–1.02) | 0.96 (0.91–1.01) | 0.94 (0.88–1.02) |
| 11 | 0.98 (0.94–1.01) | 0.96 (0.91–1.01) | 0.94 (0.87–1.01) |
| 12 | 0.98 (0.94–1.01) | 0.96 (0.91–1.01) | 0.94 (0.87–1.01) |
| 13 | 0.98 (0.94–1.01) | 0.96 (0.91–1.01) | 0.94 (0.87–1.01) |
| 14 | 0.98 (0.95–1.01) | 0.96 (0.92–1.01) | 0.94 (0.88–1.01) |
| 15 | 0.98 (0.95–1.01) | 0.97 (0.93–1.01) | 0.95 (0.89–1.02) |
| 16 | 0.99 (0.96–1.02) | 0.98 (0.94–1.02) | 0.97 (0.91–1.03) |
| 17 | 1 (0.97–1.03) | 0.99 (0.94–1.04) | 0.98 (0.92–1.05) |
| 18 | 1.01 (0.97–1.04) | 1 (0.95–1.06) | 1 (0.93–1.08) |
| 19 | 1.02 (0.97–1.06) | 1.02 (0.95–1.09) | 1.02 (0.93–1.12) |
| 20 | 1.03 (0.97–1.08) | 1.03 (0.95–1.12) | 1.04 (0.93–1.17) |
| 21 | 1.04 (0.97–1.11) | 1.05 (0.95–1.16) | 1.07 (0.93–1.22) |

Note: *Significant Increase; †Significant decrease

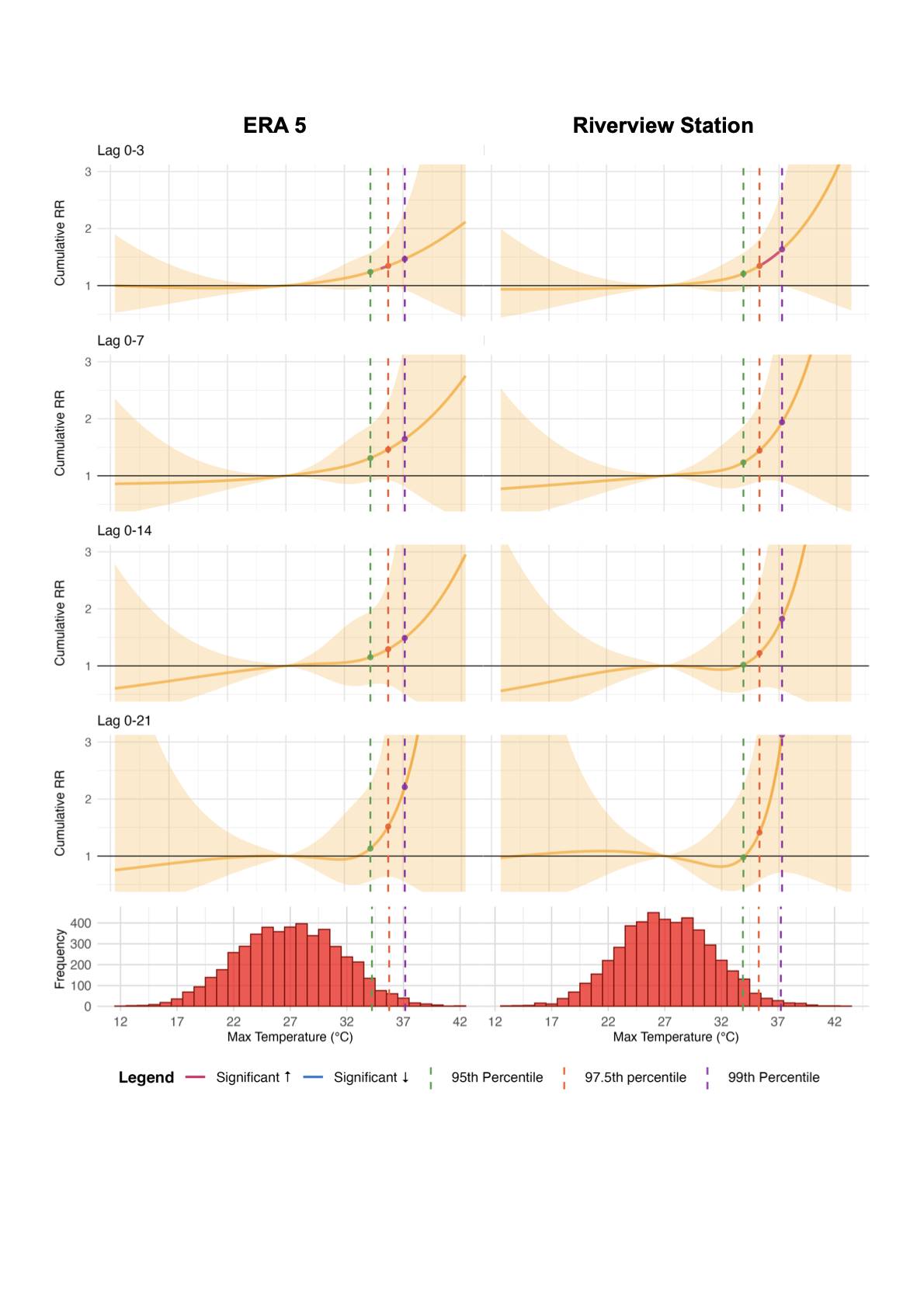
Figure S1. Cumulative exposure-response curves of maximum daily temperature (°C), based on ERA5 data, and cumulative relative risk of admission for mental disorder (relative to the median temperature). Orange, red, and blue response curve areas correspond to non-significant, significant increased, and significant decreased relative risk (RR). Green, dark orange, and purple dotted vertical lines correspond to 95th, 97·5th and 99th temperature percentile. Shaded areas correspond to 95% confidence intervals. Bottom: Histograms on daily maximum temperatures 2011-2023, with 95th, 97·5th and 99th temperature percentiles.

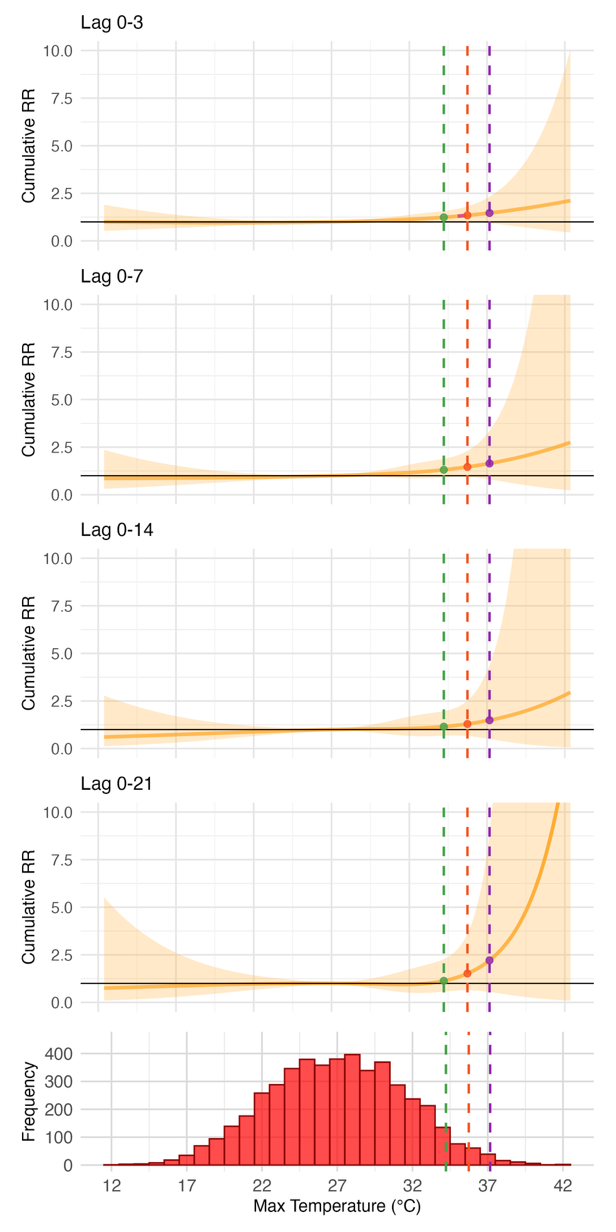
Figure S2. Cumulative (left) and lag-specific (right) exposure-response curves for maximum daily temperature (°C) and relative risk of admission for mental disorder (relative to the median temperature) for all ERA5 data

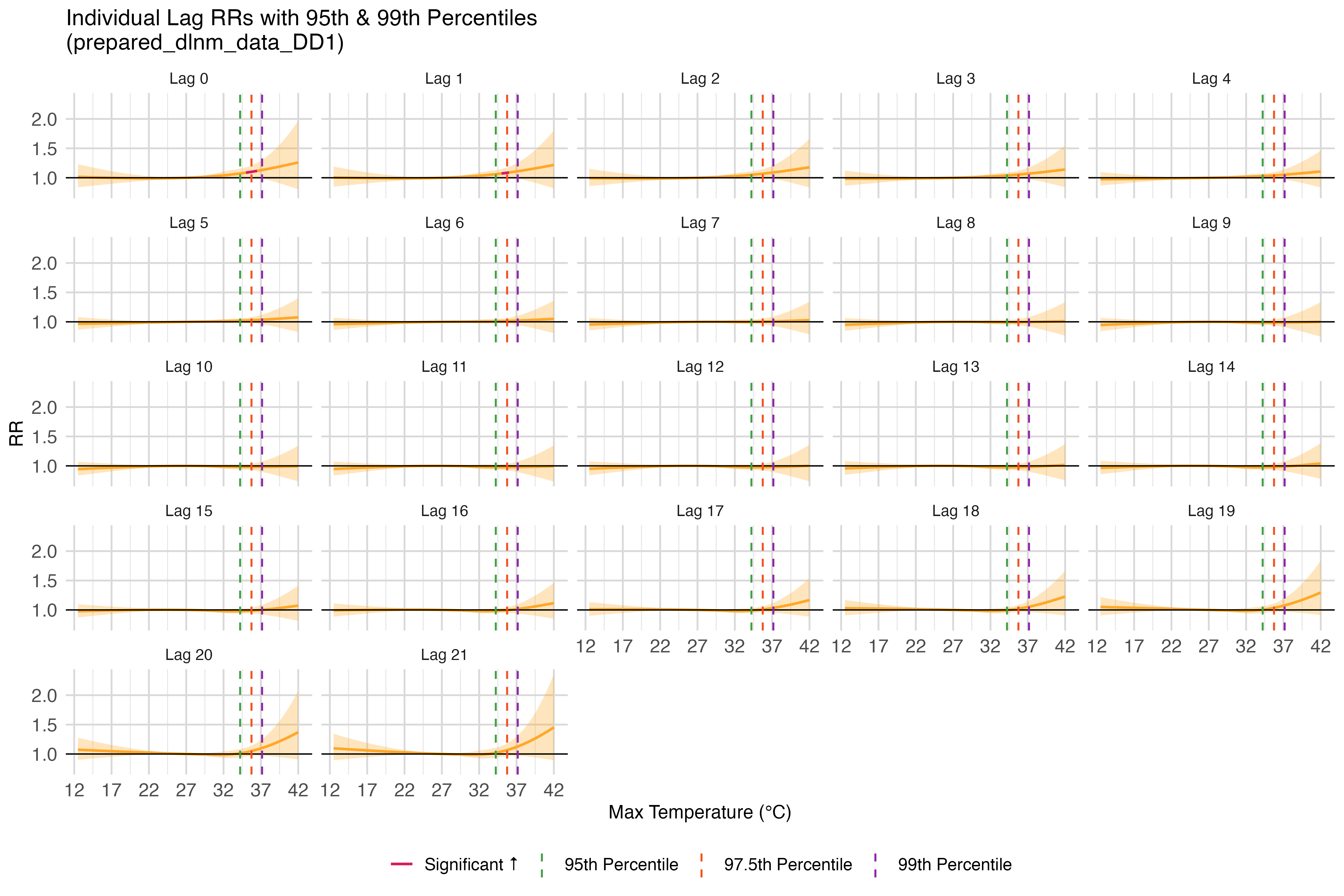

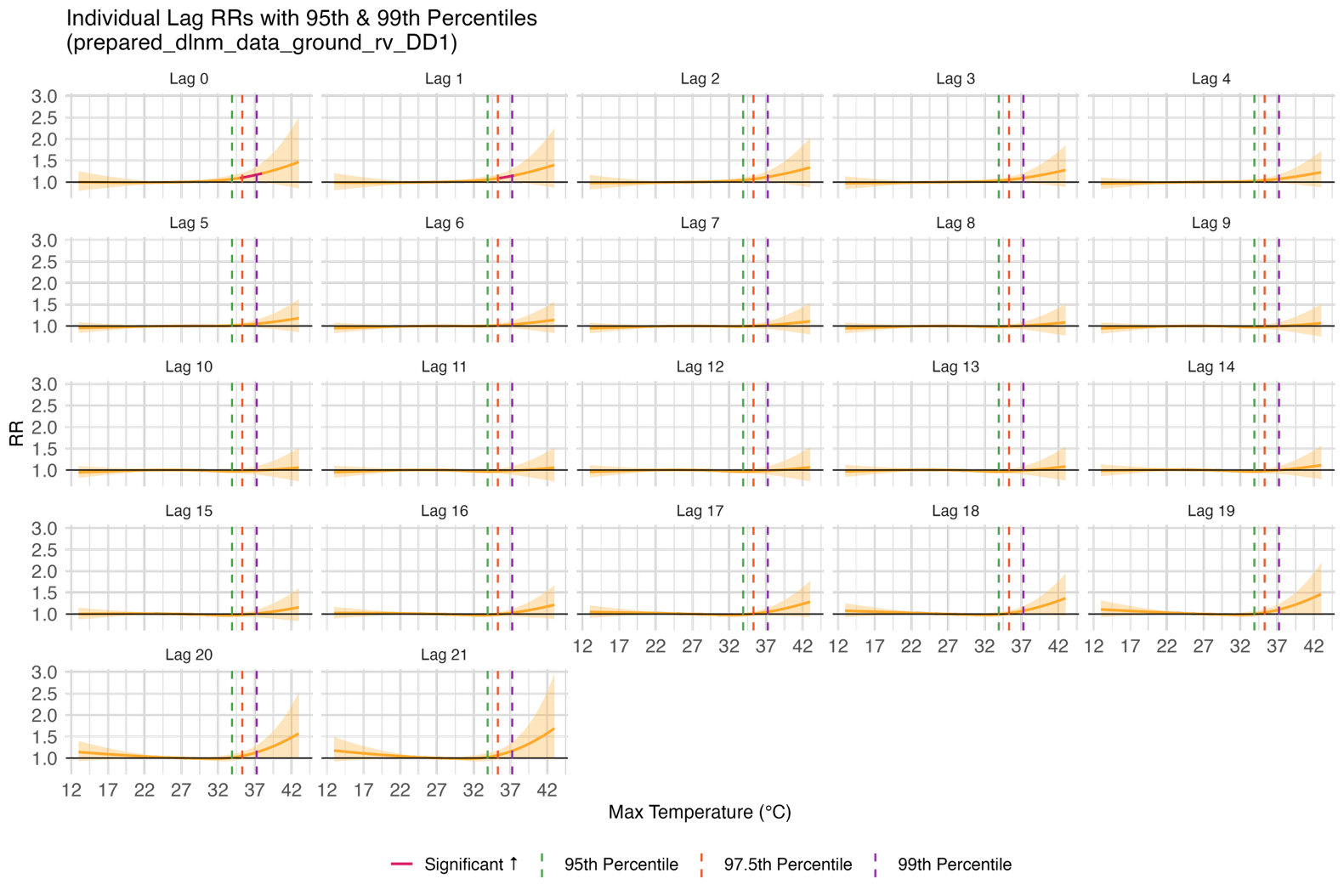

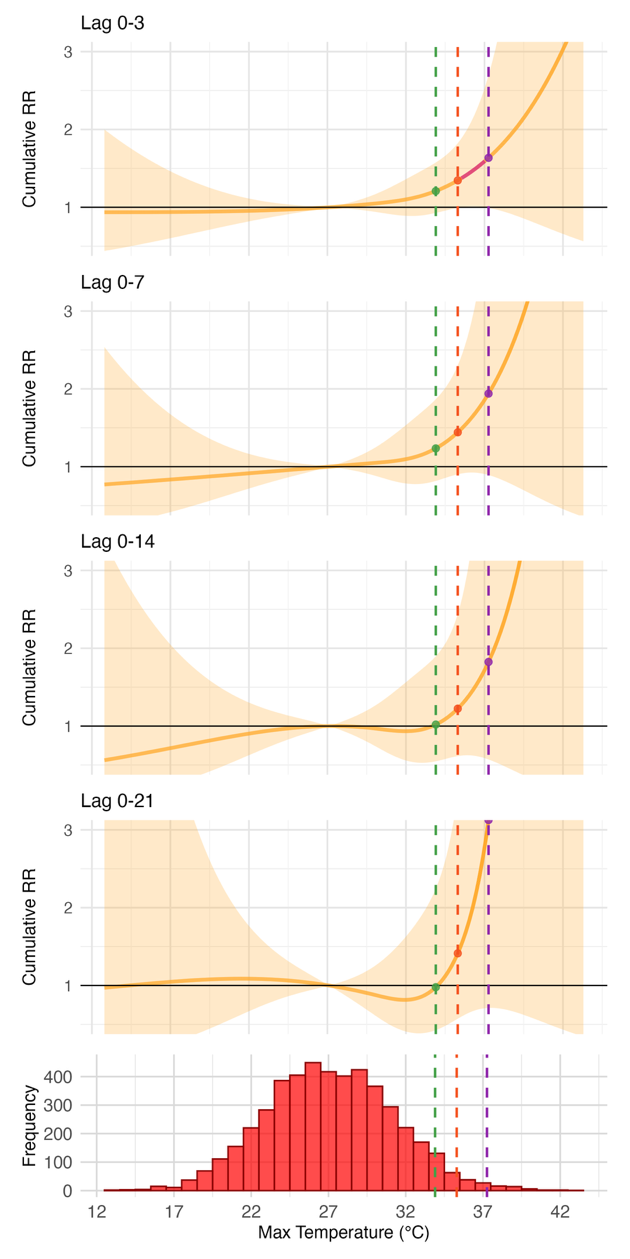
Figure S3. Cumulative (left) and lag-specific (right) exposure-response curves for maximum daily temperature (°C) and relative risk of admission for mental disorder (relative to the median temperature) for all Riverview station data

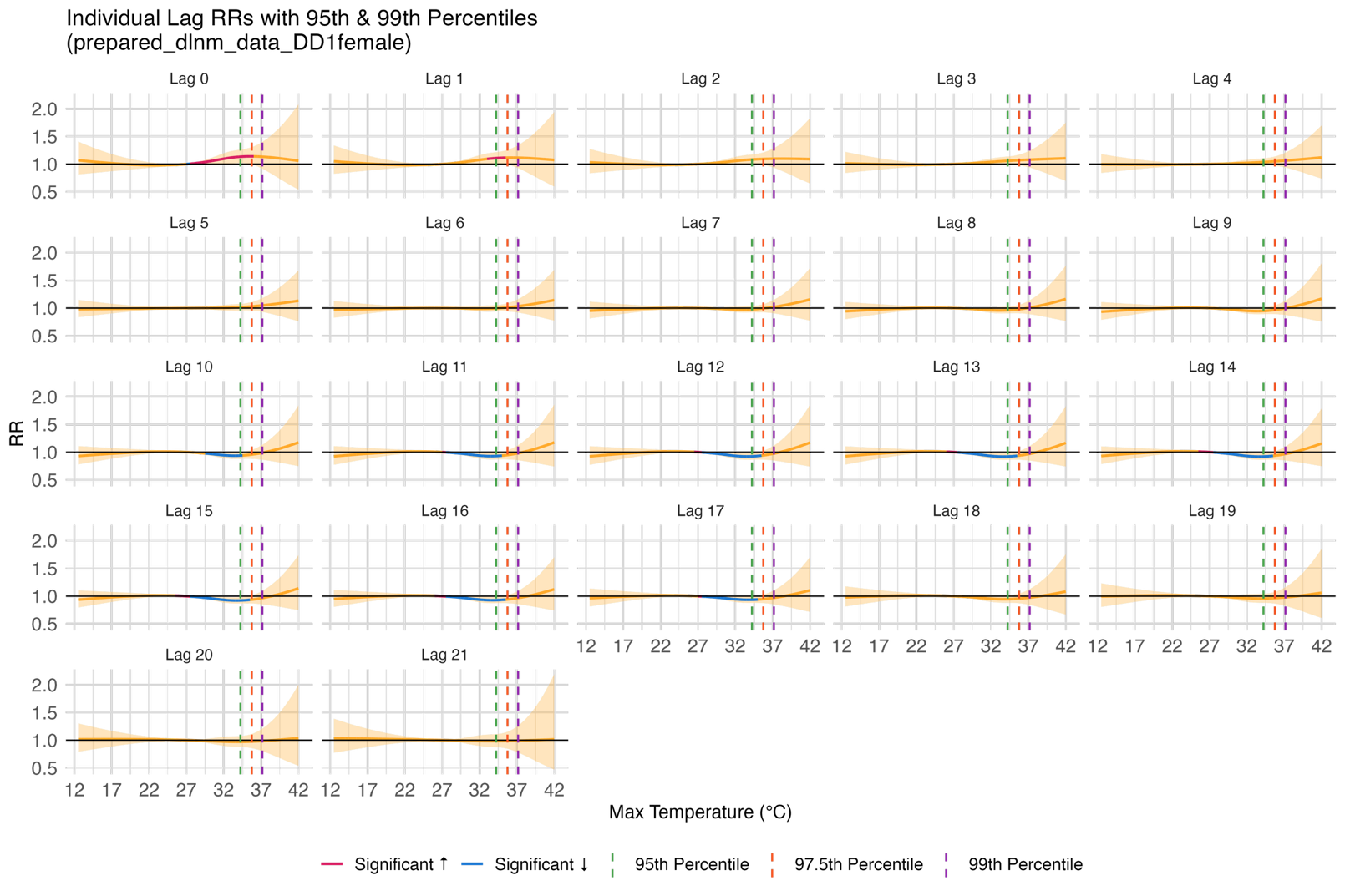

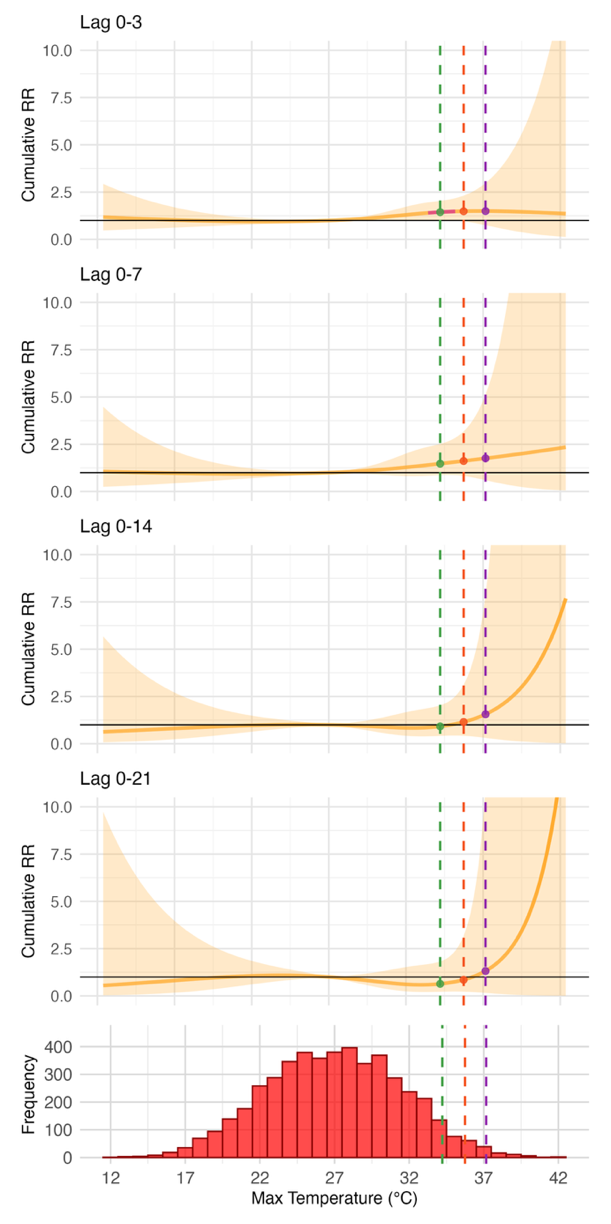

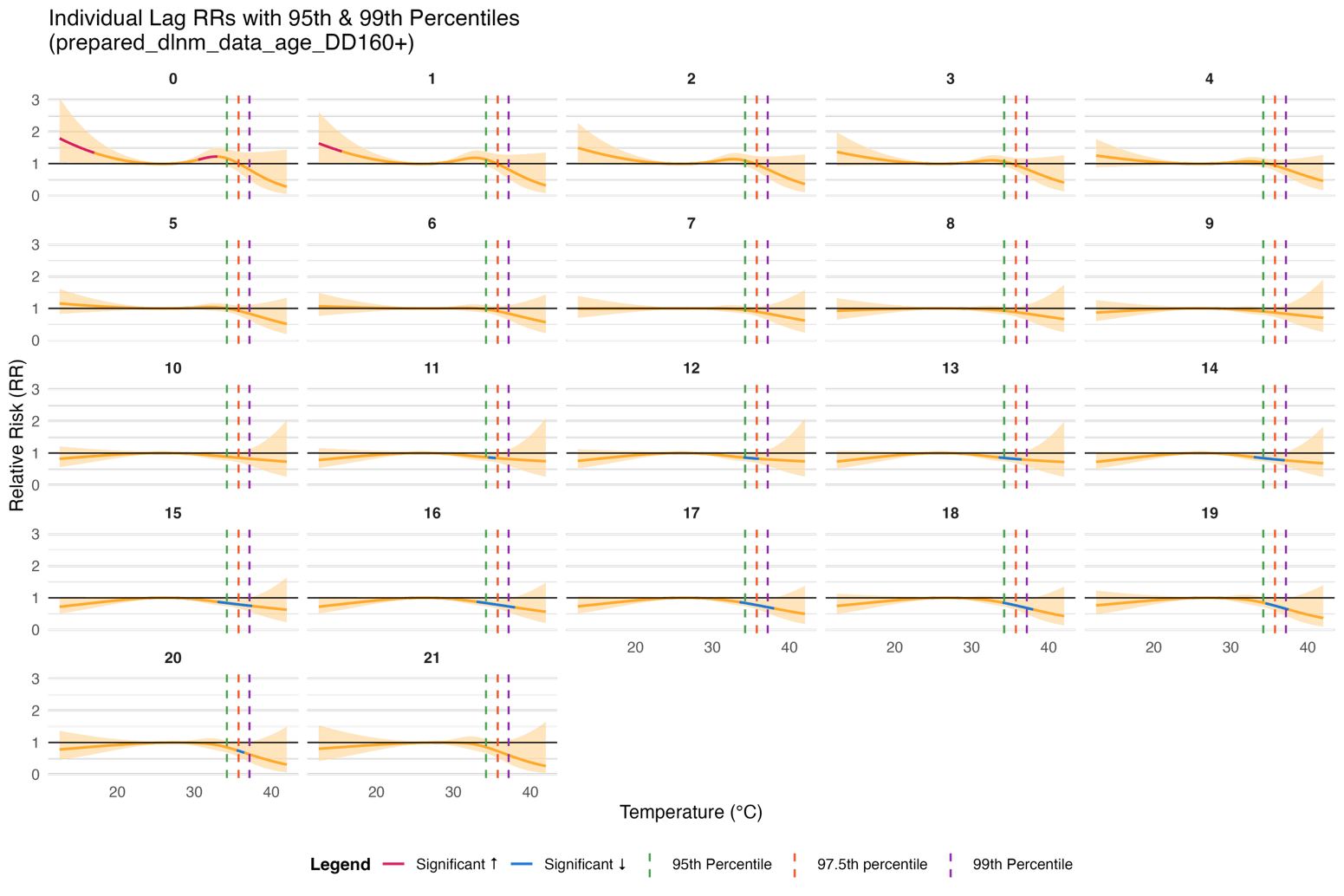
Figure S4. Cumulative (left) and lag-specific (right) exposure-response curves for maximum daily temperature (°C) and relative risk of admission for mental disorder (relative to the median temperature) for females

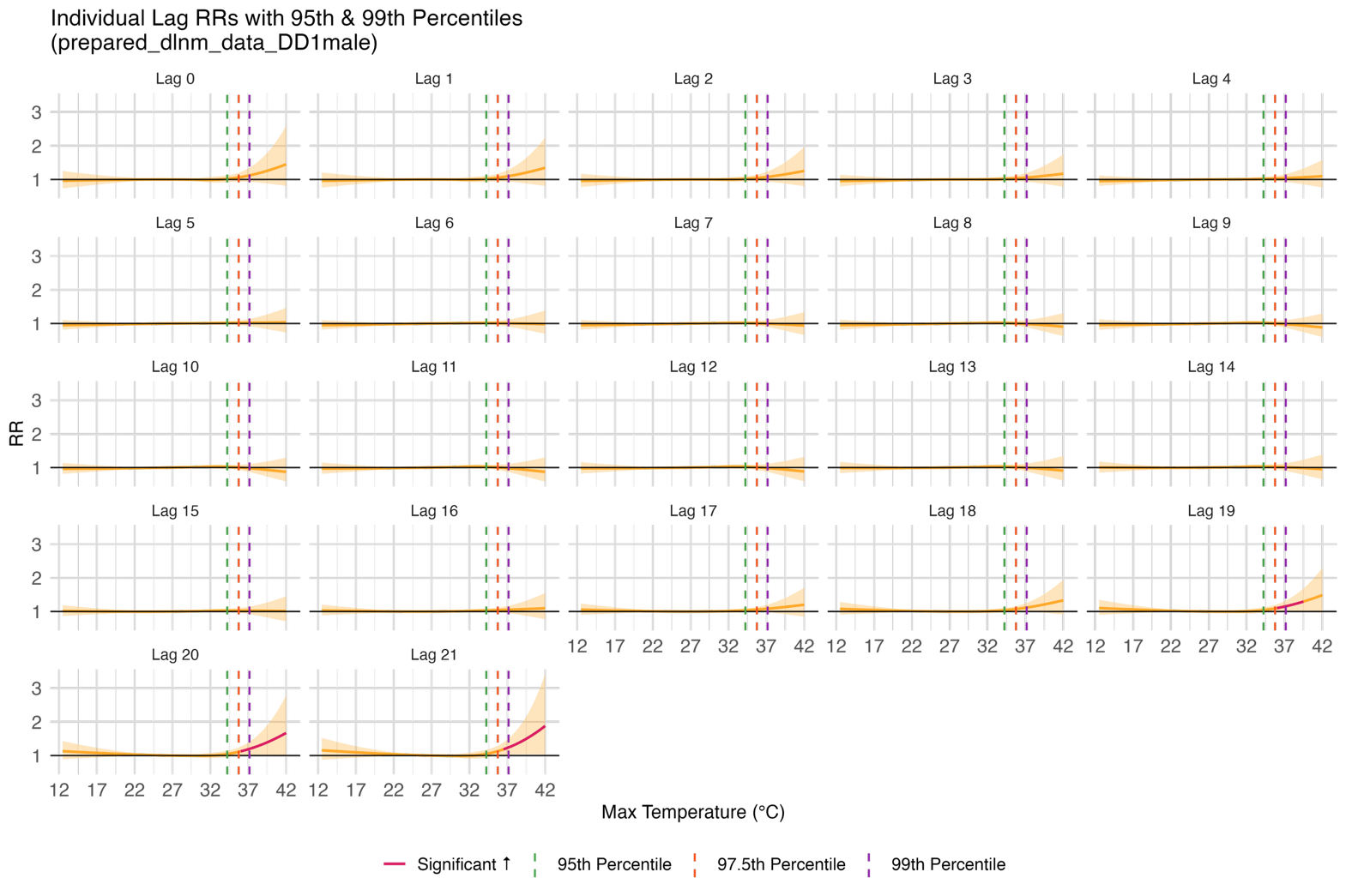

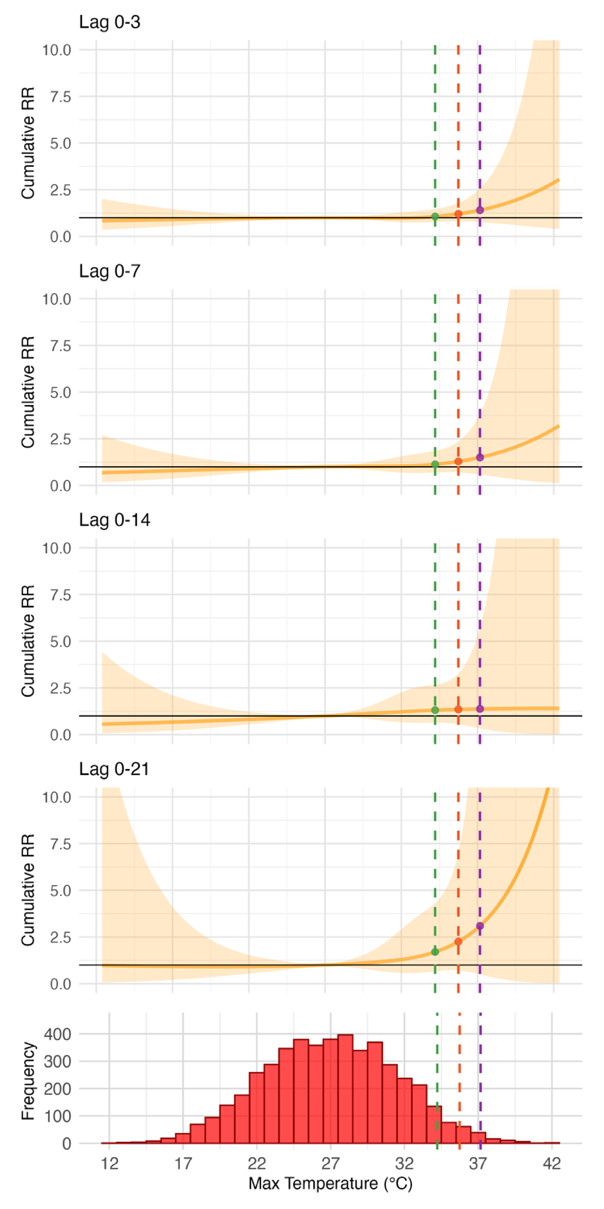

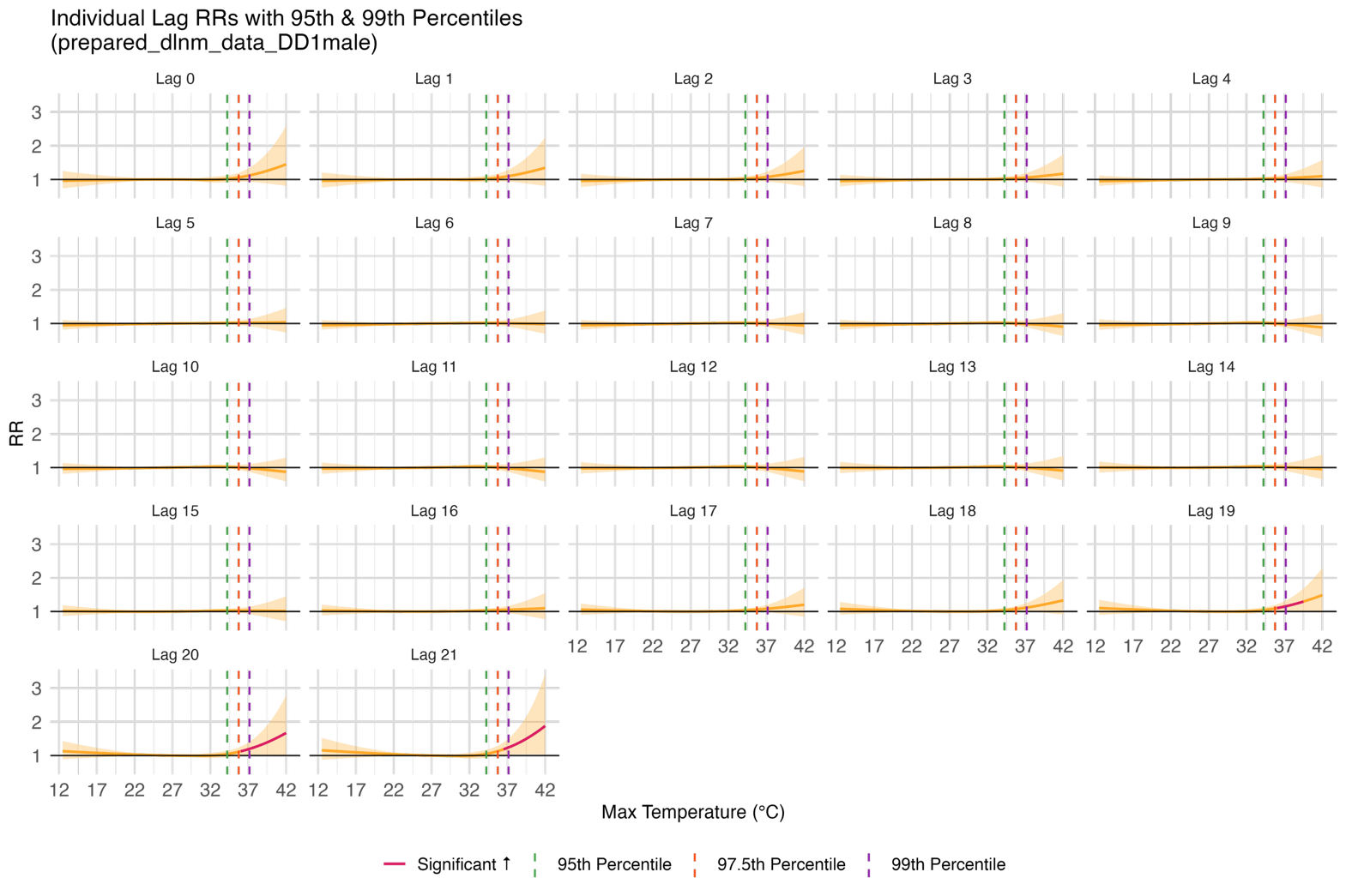
Figure S5. Cumulative (left) and lag-specific (right) exposure-response curves for maximum daily temperature (°C) and relative risk of admission for mental disorder (relative to the median temperature) for males

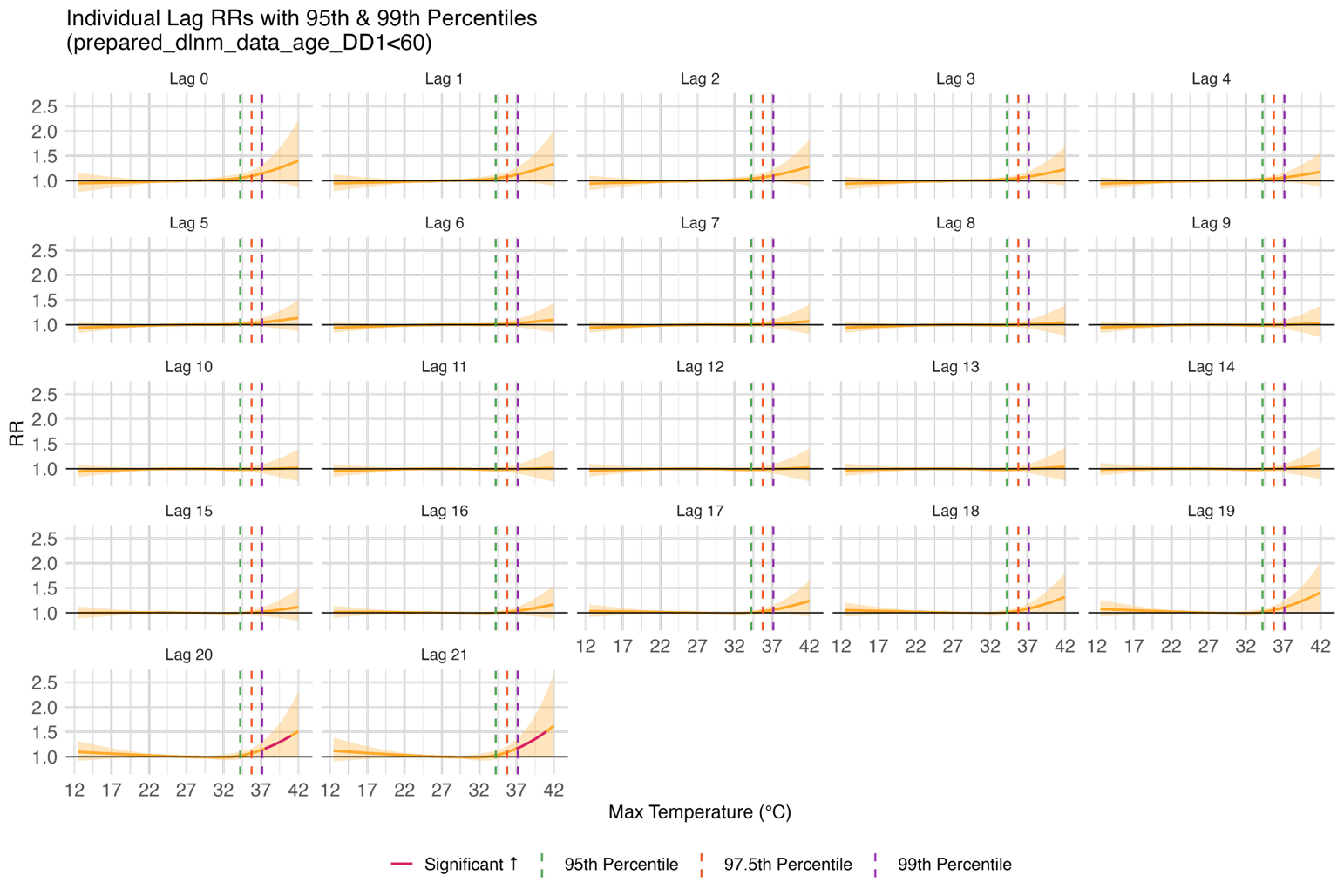

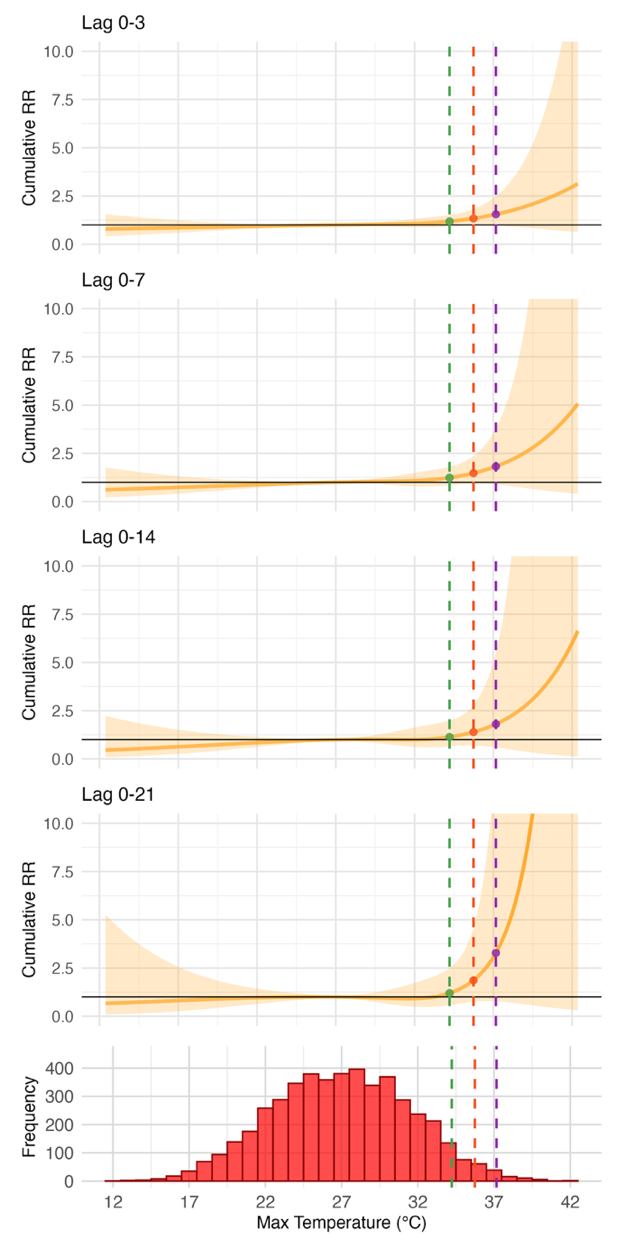

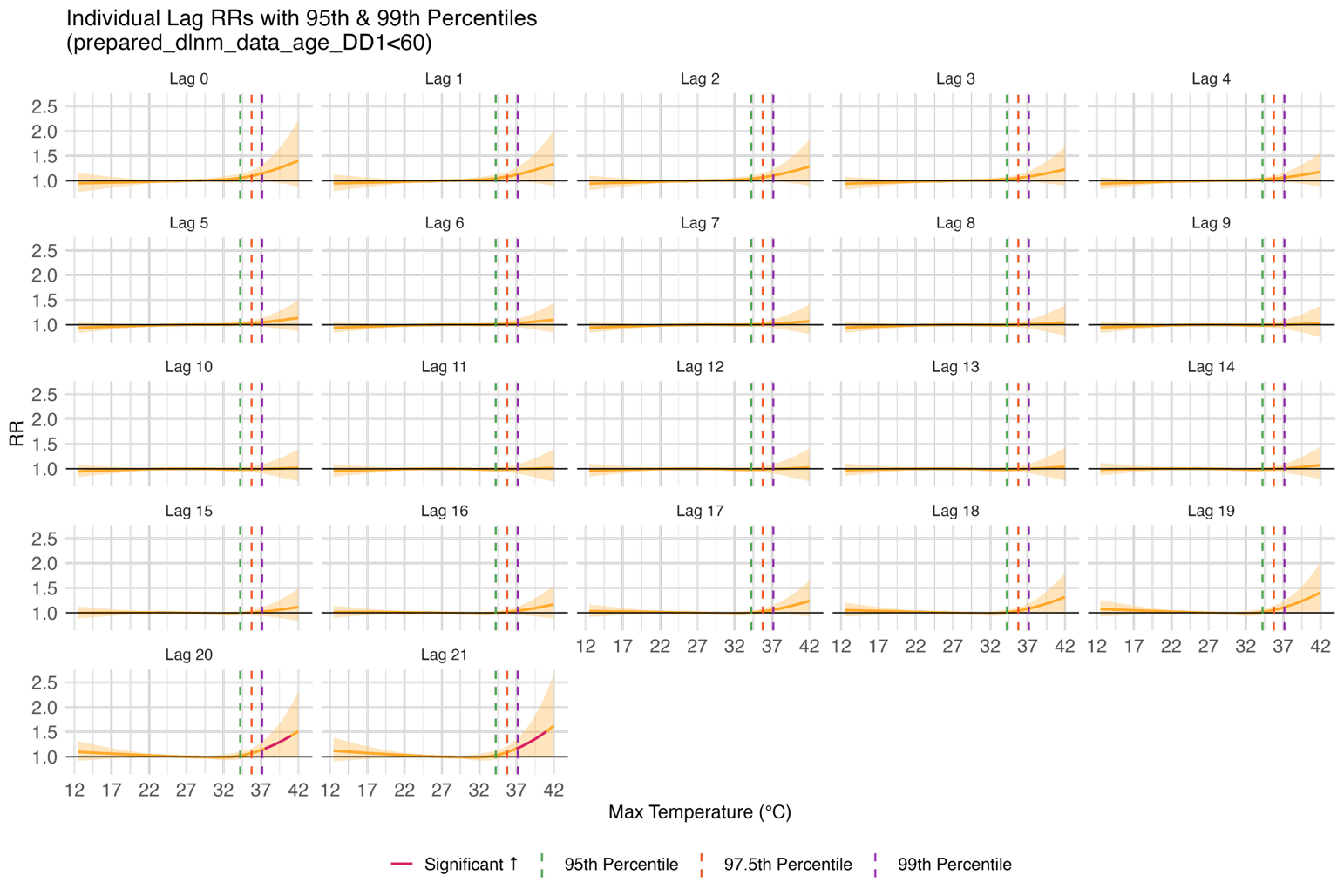
Figure S6. Cumulative (left) and lag-specific (right) exposure-response curves for maximum daily temperature (°C) and relative risk of admission for mental disorder (relative to the median temperature) for age <60

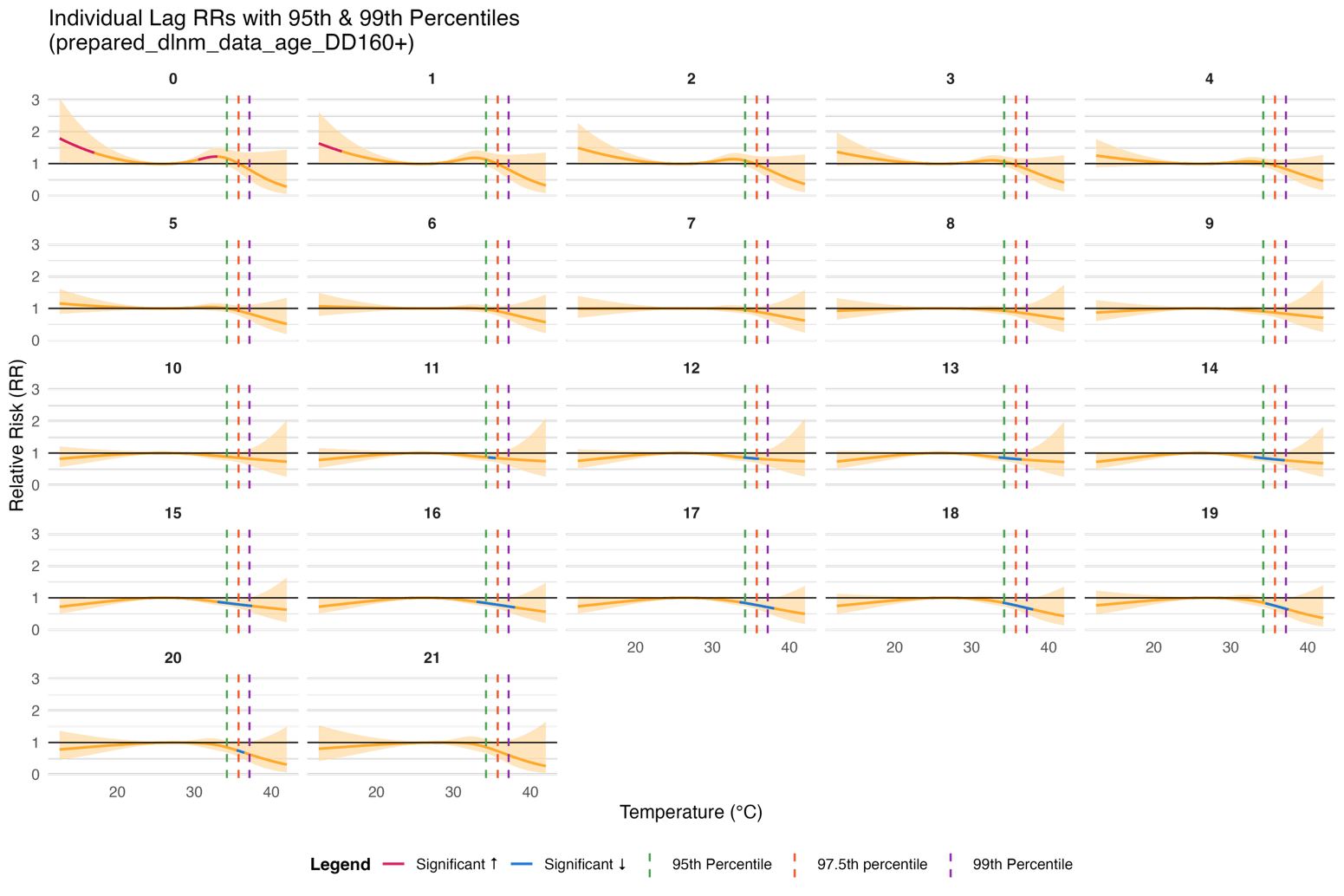

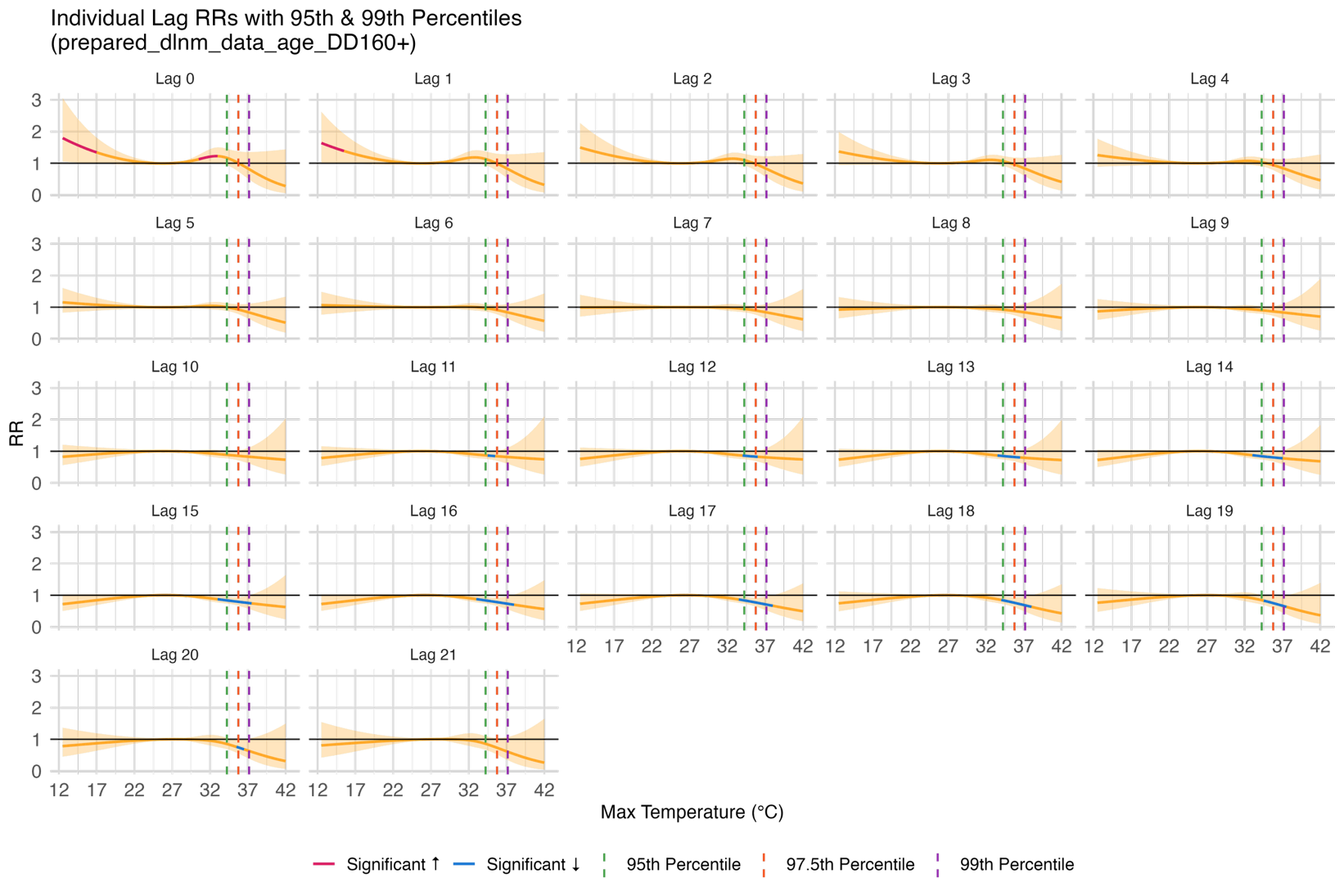

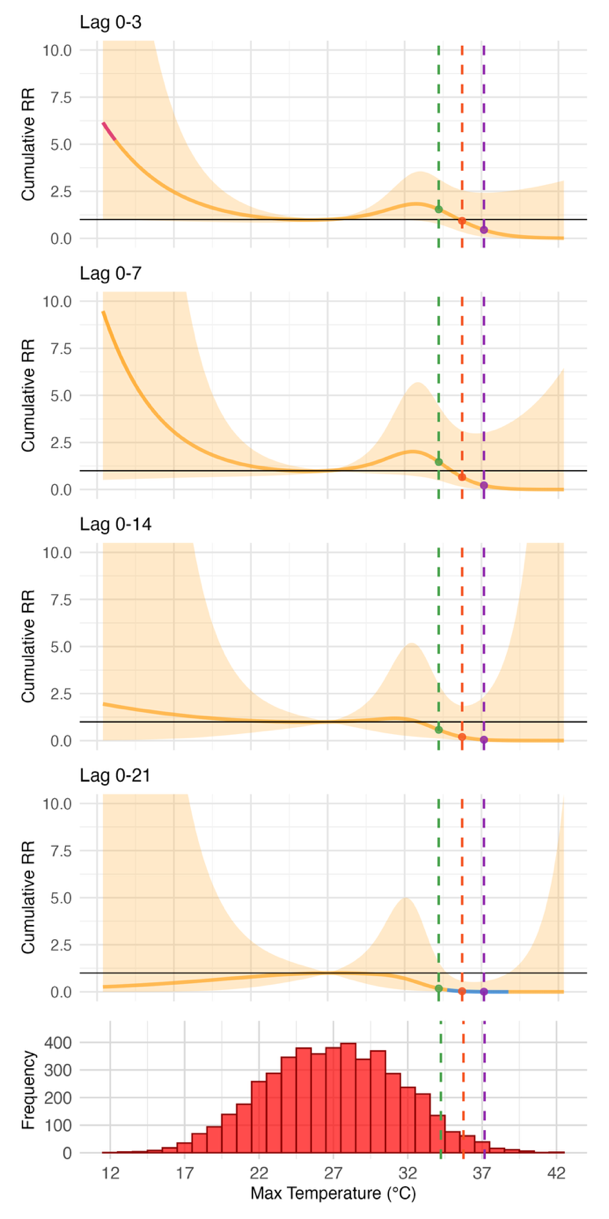
Figure S7. Cumulative (left) and lag-specific (right) exposure-response curves for maximum daily temperature (°C) and relative risk of admission for mental disorder (relative to the median temperature) for individuals age ≥60

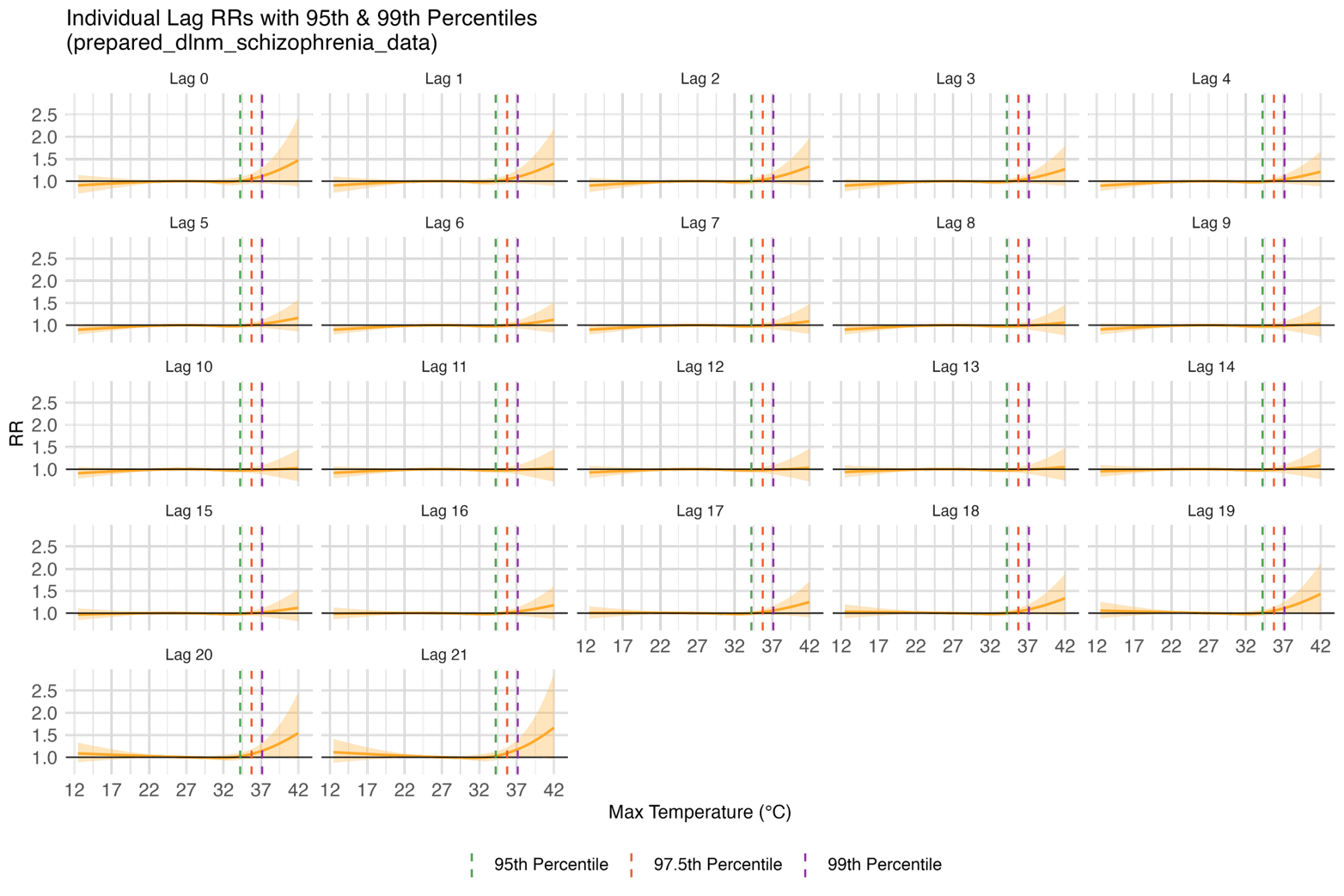

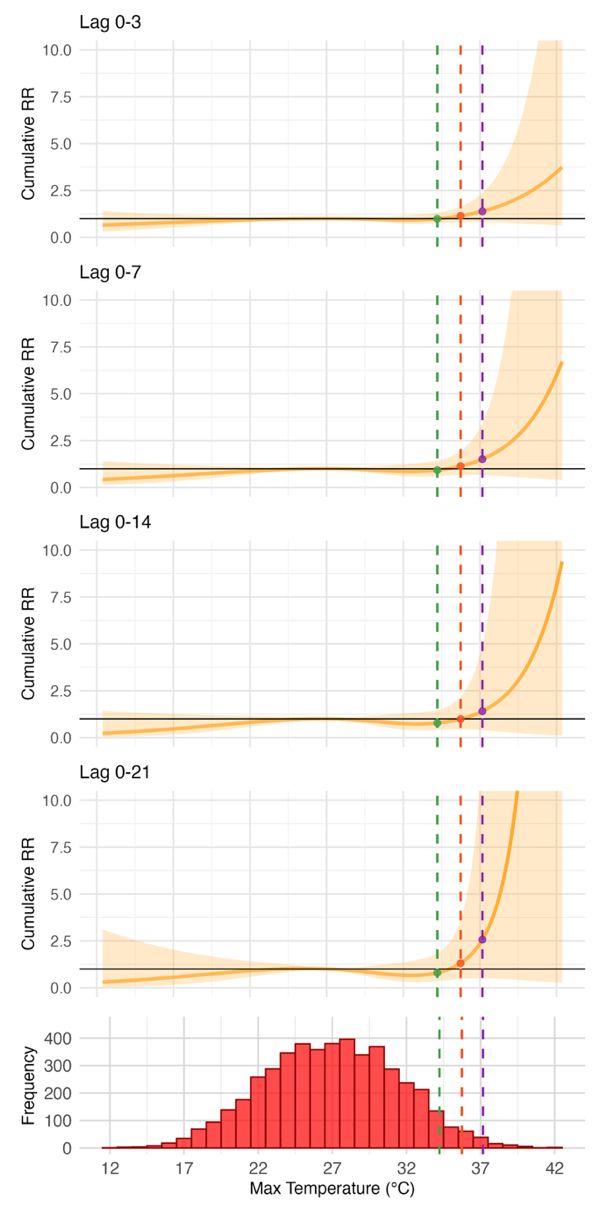

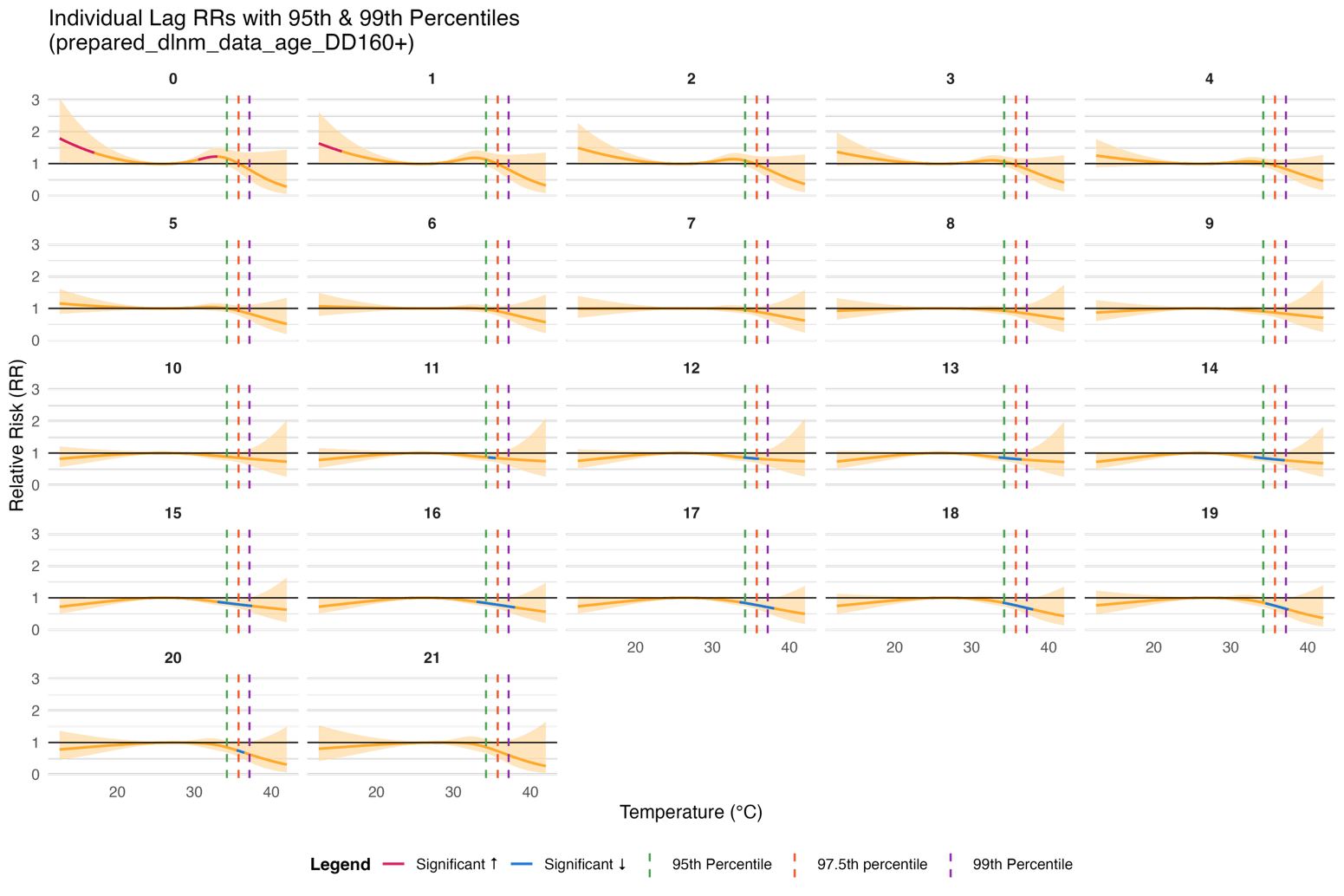
Figure S8. Cumulative (left) and lag-specific (right) exposure-response curves for maximum daily temperature (°C) and relative risk of admission for mental disorder (relative to the median temperature) for individuals diagnosed with schizophrenia

Figure S9. Cumulative (left) and lag-specific (right) exposure-response curves for maximum daily temperature (°C) and relative risk of admission for mental disorder (relative to the median temperature) for individuals diagnosed with mental health diagnoses other than schizophrenia

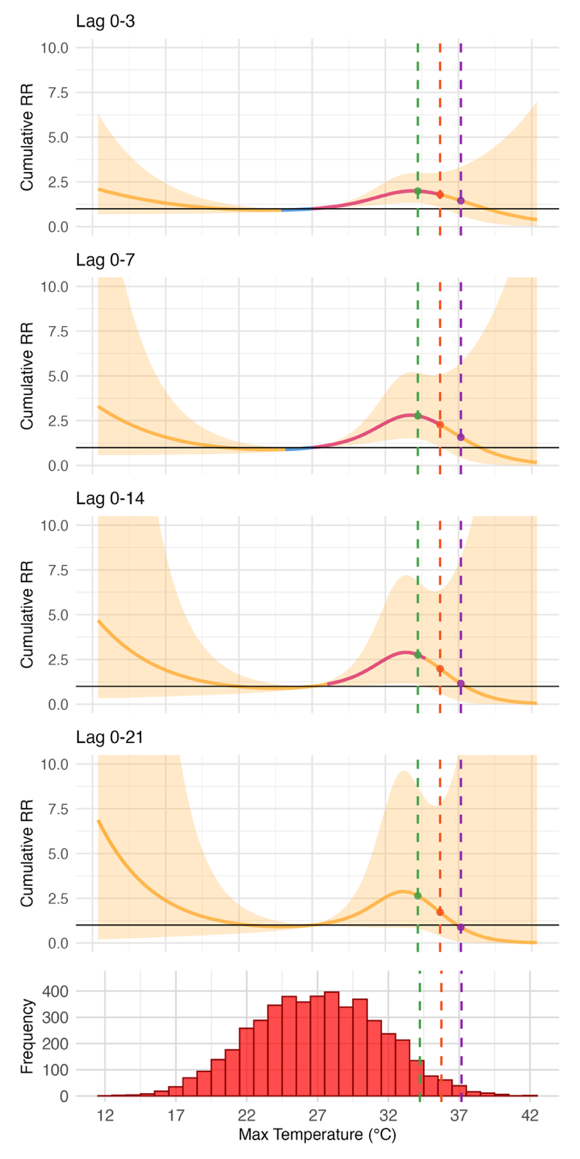

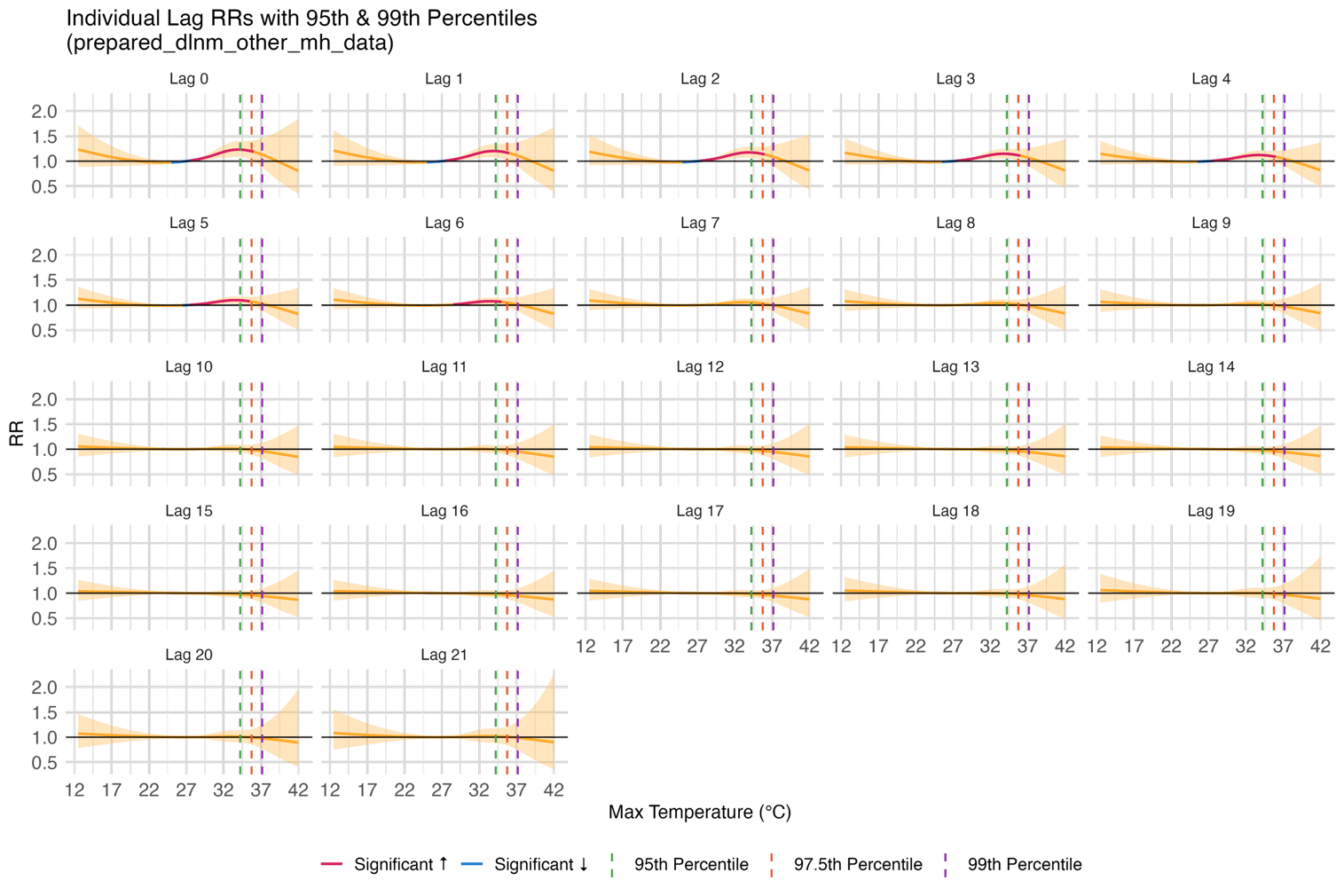

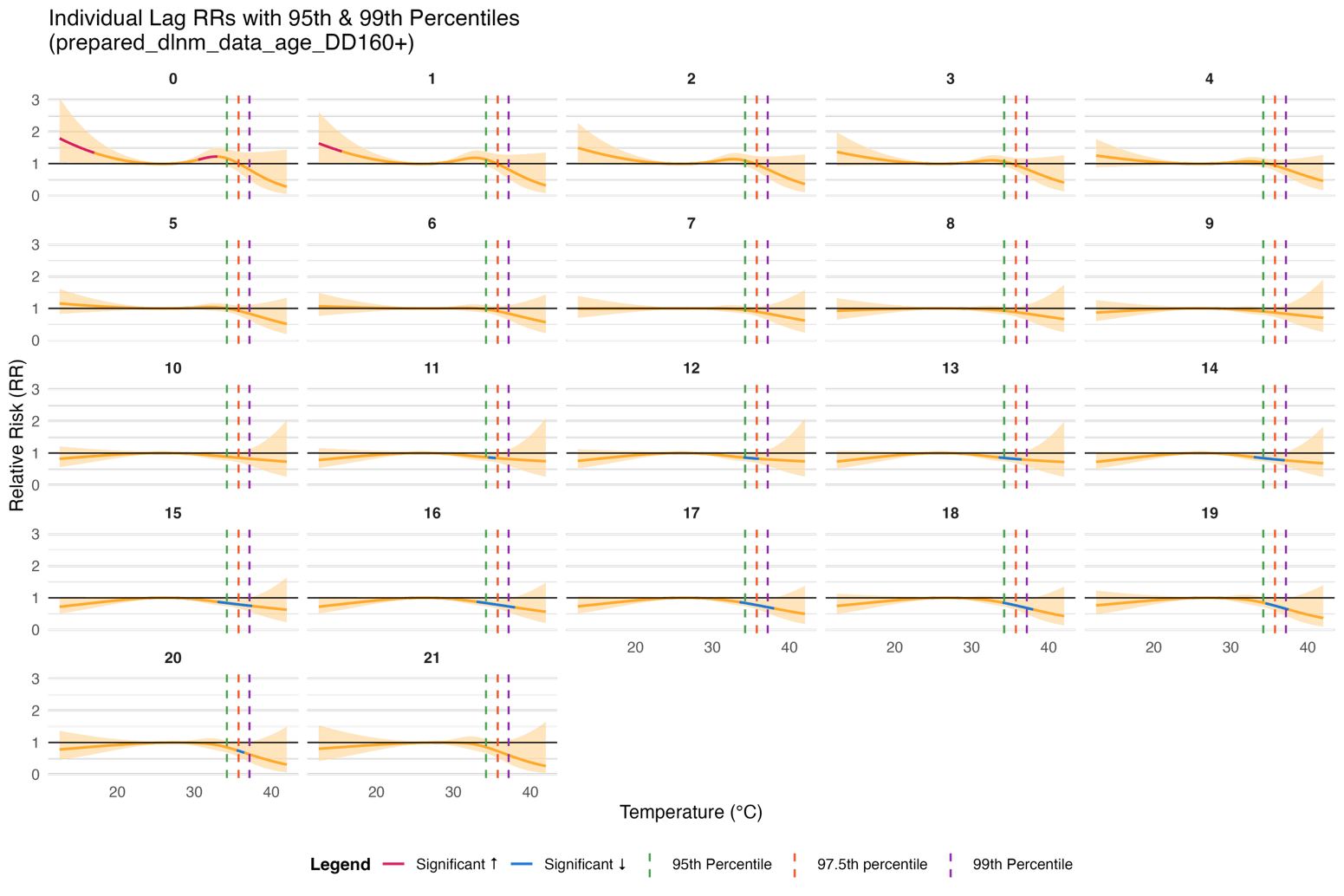

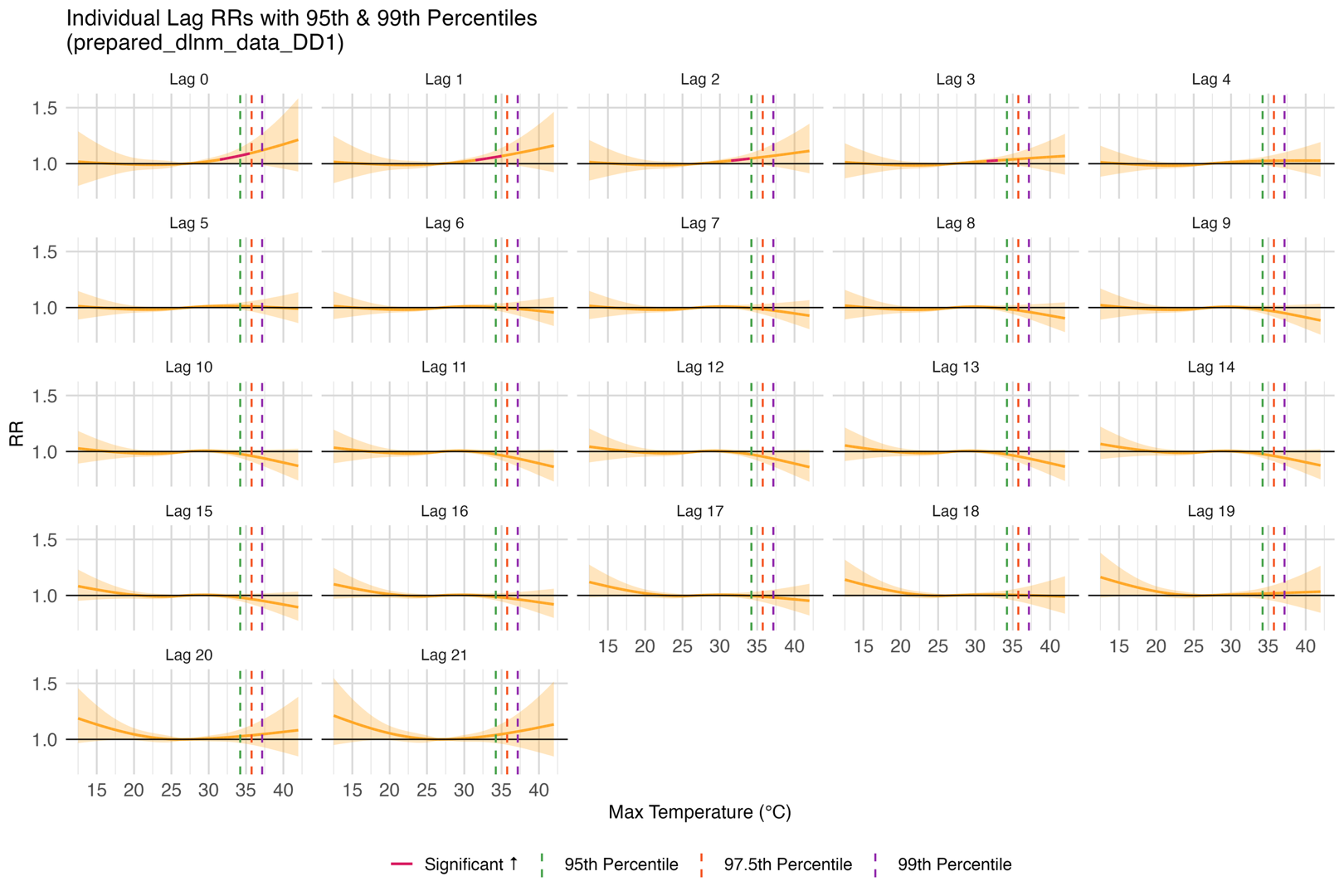

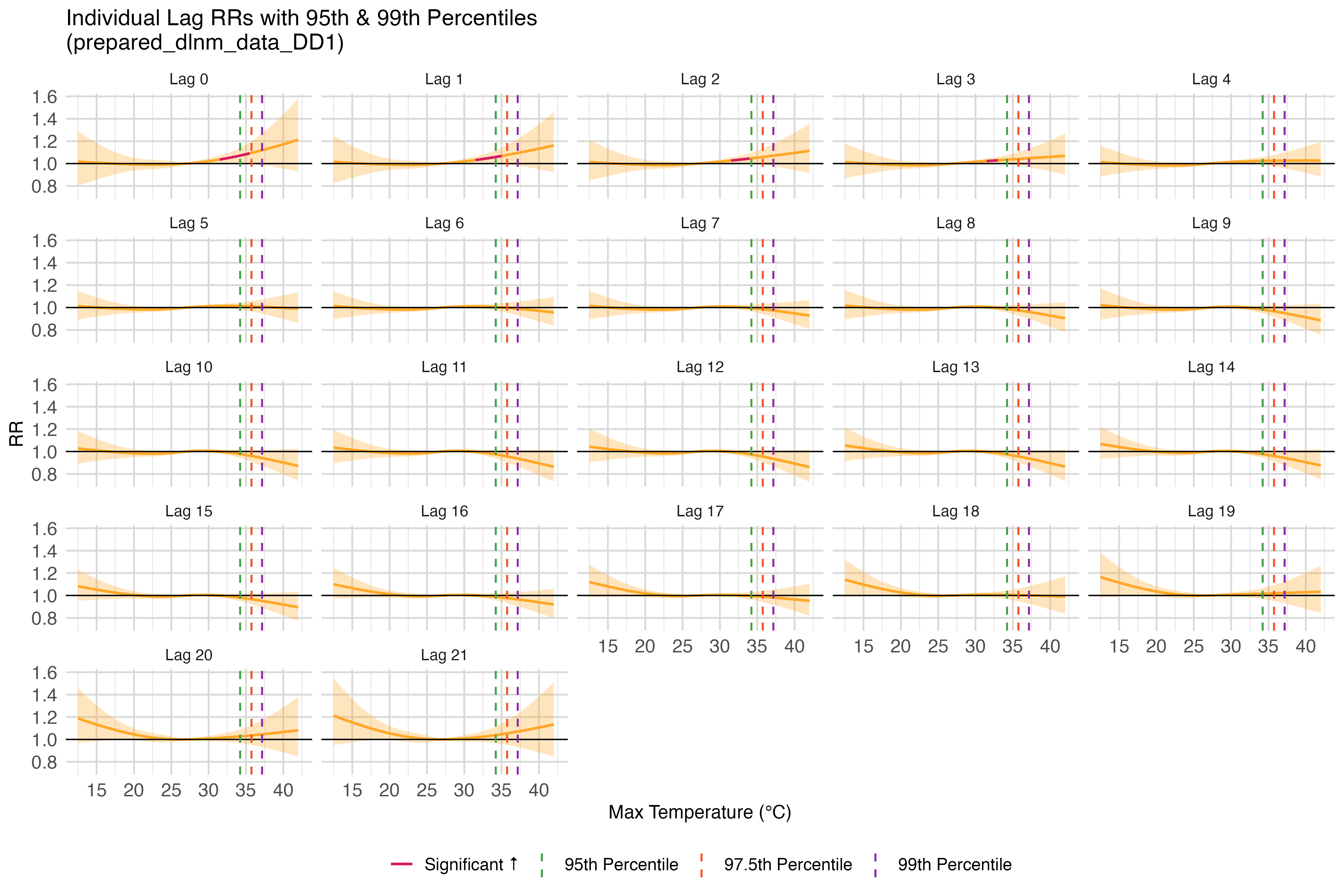

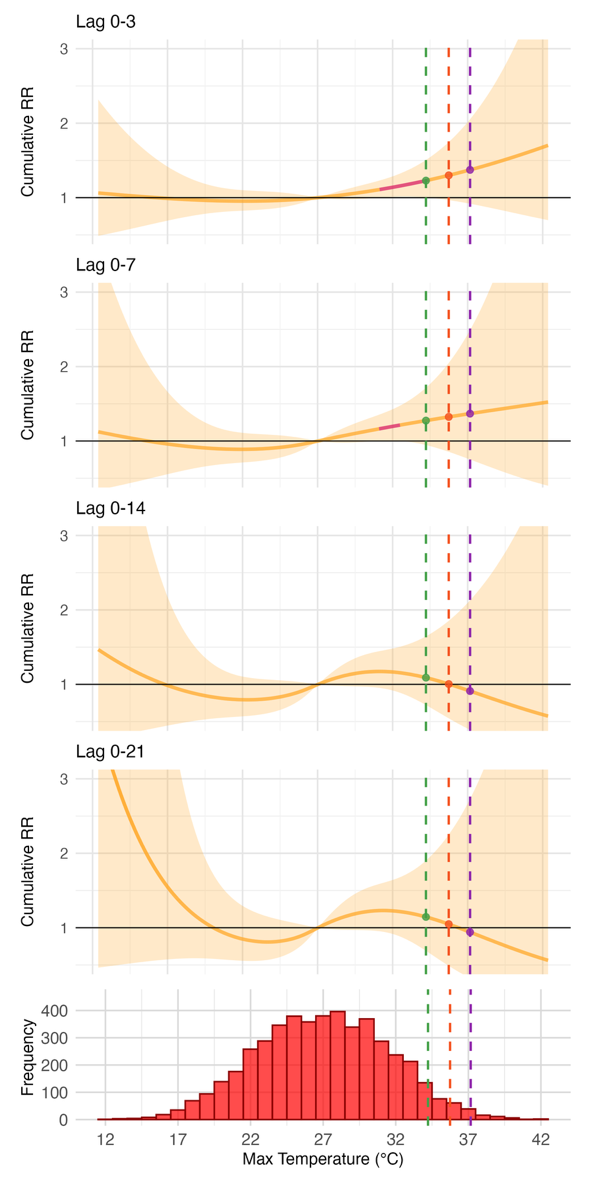
Figure S10. Cumulative (left) and lag-specific (right) exposure-response curves for maximum daily temperature (°C) and relative risk of admission for mental disorder (relative to the median temperature) for all ERA5 data (sensitivity analysis **-** seasonality degrees of freedom optimized)
